# Brain functional dynamics linked to depression and anxiety: cross-sectional and longitudinal associations over 9 years

**DOI:** 10.64898/2026.09.08.26362586

**Authors:** Julian Gaviria Lopez, Guido Van Wingen, Rick Jansen, Jennifer Labus, Brenda W.J.H. Penninx

**Affiliations:** Psychiatry department, Amsterdam UMC Location Vrije Universiteit, Amsterdam, the Netherlands; Amsterdam Neuroscience, Brain Imaging, Amsterdam, the Netherlands. Anatomy and Neurosciences, De Boelelaan 1117, Amsterdam, The Netherlands; Amsterdam Neuroscience, Mood, Anxiety, Psychosis, Sleep & Stress Program, Amsterdam, the Netherlands; G. Oppenheimer Center for Neurobiology of Stress and Resilience, Vatche & Tamar Manoukian Division of Digestive Diseases, Department of Medicine, David Geffen School of Medicine, University of California, Los Angeles, California

**Author notes:** **Correspondence:** Julian Gaviria Lopez.

**Keywords:** Naturalistic course of depression and anxiety, Brain networks, affective disorders, dynamic functional connectivity (dFC), depression, anxiety

## Abstract

**Background:** Resting-state fMRI (rs-fMRI) co-activation patterns (CAPs) capture recurrent whole-brain states that may index neurobiological substrates of affective psychopathology, but whether their temporal dynamics co-evolve with symptom severity over the naturalistic long-term course of depression and anxiety remains unknown.

**Methods:** We examined rs-fMRI and clinical data from 340 participants of the NESDA neuroimaging cohort [228 with major depression (MDD) and/or an anxiety disorder (AFF), 112 controls (CTR)] collected at baseline (Y0), 2-year (Y2), and 9-year (Y9) follow-up. Four CAPs were identified, internally validated, externally replicated against an independent dataset, and quantified by dwell time (time spent of each brain CAP) and entries (initiation of each CAP). These metrics were related to depression symptomatology (IDS score), anxiety (BAI), and fear (FQ) severity at two levels: cross-sectionally, between subjects and groups, and longitudinally, through within-person associations between changes in CAP expression and changes in symptoms severity.

**Results:** At baseline, attention–somatomotor (AT-SM_CAP_) dwell time was negatively associated with depression severity in controls (rho = −0.29, *q* < 0.05), an association absent in individuals with AFF ( rho = 0.09, *q* = 0.79;). Longitudinally, group-level CAP trajectories did not differ between AFF and CTR, but within-person changes in frontoparietal–default-mode (FP-DM_CAP_) dwell time tracked changes in depression severity exclusively in AFF (Y2→Y9: rho = 0.47, *q* = 0.04). Patients whose FP-DM_CAP_ dwell time increased were those whose depression failed to improve.

**Conclusions:** CAP dwell time carried clinical meaning at two levels: AT-SM_CAP_ distinguished health from illness at baseline, while FP-DM_CAP_ dwell time indexed within-person illness course over nine years. Because FP-DM_CAP_ co-activates frontoparietal control with default-mode regions implicated in rumination, its rebound in dwell time may reflect a shift toward perseverative self-referential processing in patients who fail to remit. These findings position CAP dwell-time–symptom coupling as a neurobiologically grounded, individually resolved marker of naturalistic illness course relevant to precision psychiatry.

## 1. INTRODUCTION

Depression and anxiety are leading causes of disability worldwide (1). Their course is protracted and heterogeneous, with some individuals remitting, others relapsing, and many remaining chronically symptomatic over years (2,3). Despite an enormous research investment in the last decades (4,5), psychiatry still lacks robust, validated biomarkers that make diagnosis based on self-report and clinical interview more efficient (6). Self-report instruments capture symptom course, yet the neurobiological markers proposed to outperform them have largely failed to demonstrate added clinical value (4). One reason may be methodological: candidate biomarkers are typically evaluated only through cross-sectional, between-subject comparisons, establishing differences between individuals with these disorders and controls, or derived from short-interval designs, rather than through direct tests of person-specific illness trajectories over the long-term course.

This methodological gap is compounded by a further limitation specific to neuroimaging studies. Most fMRI-based studies on depression and anxiety (7) have relied on static functional connectivity (FC), which averages the BOLD signalling across the scan and imposes assumptions of temporally homogeneous coupling (8). Consequently, these FC approaches may miss the transient network reconfigurations, captured by dynamic functional connectivity (dFC), that have been implemented for a few studies in the neural characterization of affective dysfunction (9,10). Within the dFC framework, the co-activation pattern (CAP) approach offers a data-driven alternative, identifying recurring whole-brain states and their temporal dynamics at frame-wise resolution (11,12). Among the available temporal metrics, dwell time (how long a state persists once entered) has proven the most consistently informative for linking CAP dynamics to affective states in bipolar disorder (13,14), depression (15,16), schizophrenia (17), and the temporal inertia of negative emotional states and their reconfiguration after affective challenge (18). A complementary metric, the number of entries into a state (how many separate times a state is initiated over a scan, independent of how long each episode lasts), captures state-initiation frequency rather than persistence, and is examined here alongside dwell time to establish whether the two temporal properties carry distinct or overlapping clinical information.

Capturing the fluctuating course of affective disorders requires large-scale longitudinal designs with repeated clinical and biological assessments spanning years. This is precisely the infrastructure that the Netherlands Study of Depression and Anxiety (NESDA; www.nesda.nl) was built to provide: a multi-site naturalistic cohort of over 2,900 individuals with depressive and anxiety disorders and healthy controls, followed across twelve years with regular reassessment of diagnostic status, symptom severity, and biological measures (2,3). In NESDA, 67% of individuals with a depressive disorder had a current comorbid anxiety disorder, and such comorbidity consistently predicts a more chronic course (19). Therefore, we combined both diagnostic groups into a single affective (AFF) group given their shared course and high co-occurrence (20). The present study draws on a subset of NESDA (the neuroimaging cohort, N = 340) including healthy controls and participants with major depression and/or an anxiety disorder, with resting-state fMRI acquired at baseline (Y0), two-year (Y2), and nine-year follow-up (Y9). We test two complementary hypotheses. First (H1), that CAP temporal dynamics are associated with symptom severity cross-sectionally. Second (H2), that CAP temporal dynamics track the longitudinal course of depression, anxiety, and fear over the nine-year follow-up. By resolving brain–symptom coupling within individuals across nearly a decade, this study aims to establish whether the dynamics of brain connectivity provide a neurobiological index that complements self-report measures in clinical settings, consistent with the within-person focus of precision psychiatry (21,22).

## 2. METHODS

### 2.1. Participants

The NESDA study (www.nesda.nl) was approved by the ethical review boards of all participating centers (2), and all participants gave written informed consent. Of 340 NESDA cohort members with resting-state fMRI data (Figure 1), 228 with a current (6-month) DSM-IV diagnosis of MDD and/or an anxiety disorder (panic disorder, social phobia, generalized anxiety disorder, or agoraphobia) at baseline— established using the CIDI—constituted the affective group (AFF; 153 female; mean age 38.7 ± 11.1 years). The remaining 112 participants without these diagnoses formed the control group (CTR; 69 female; mean age 42.5 ± 10.7 years. Table 1).

**Figure 1.**
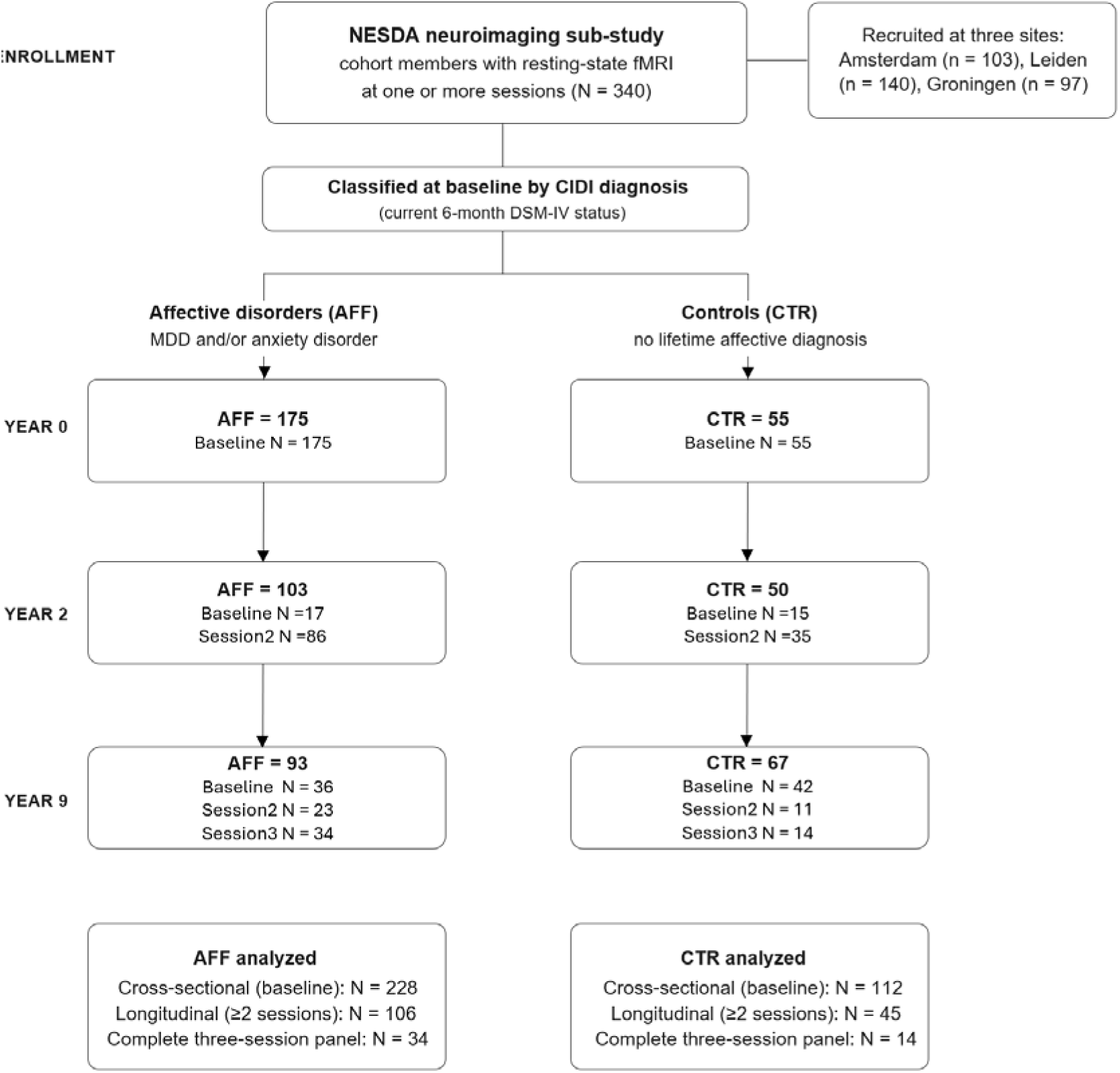
CONSORT flow diagram for the NESDA neuroimaging sub-study. Flow of the 340 participants (228 affective disorders [AFF]; 112 controls [CTR]) contributing usable resting-state fMRI at one or more of the three assessment sessions. Group membership was fixed by baseline diagnostic status (CIDI; current 6-month DSM-IV). Because NESDA acquired neuroimaging on a rolling basis, “baseline” denotes each participant’s earliest available session rather than a fixed calendar timepoint: within each session box, participants are partitioned by whether that session was their first (baseline), second, or third scanning session. Thus each calendar session (Year 0, Year 2, Year 9) mixes newly entering participants with returning ones, and the three boxes do not form a strict attrition cascade. Summing the baseline entries across the three sessions recovers the full cross-sectional sample. The diagram shows the number of participants with fMRI data. Counts for complete data (fMRI plus symptom scores) are reported per analysis (see Figure 4 for cross-sectional baseline counts and Figure 5 for longitudinal counts). Recruitment-site distribution and full demographic, medication, and clinical characteristics are reported in Table 1.

**Table 1.** Demographic and Clinical Characteristics. NESDA fMRI subsample. Data are presented as mean (SD) or n (%). AFF = patients with affective disorders (MDD and/or ANX). CTR = healthy controls. Group reflects current diagnostic status at each session (time-varying). Demographics and medication are reported only at y0 (baseline). Symptom scores are z-standardized across all participants and sessions. N at y2 and y9 reflects all subjects available at that session with AFF or CTR status. Higher education = university or higher vocational degree.

| Characteristic | AFF (N=228) | CTR (N=112) | Statistic | p |
| --- | --- | --- | --- | --- |
| Demographics |  |  |  |  |
| Age, years | 38.66 (11.13) | 42.46 (10.65) | t = -3.00 | 0.003 |
| Female, n (%) | 153 (67%) | 69 (61%) | χ² = 0.77 | 0.379 |
| Higher education, n (%) | 78 (34%) | 66 (58%) | χ² = 19.45 | <.001 |
| Scan site N(%) |  |  |  |  |
| Amsterdam | 61 (26%) | 33 (29%) | χ² = 2.03 | 0.362 |
| Leiden | 95 (41%) | 52 (46%) |  |  |
| Groningen | 72 (31%) | 27 (24%) |  |  |
| Medication N(%) |  |  |  |  |
| SSRI | 59 (25%) | 3 (2%) | - | - |
| Benzodiazepine | 19 (8%) | 1 (0%) | - | - |
| Symptom severity |  |  |  |  |
| Depression (IDS) | 0.68 (0.83) | -0.94 (0.41) | t = 19.48 | <.001 |
| Anxiety (BAI) | 0.58 (1.04) | -0.76 (0.44) | t = 12.96 | <.001 |
| Fear (FQ) | 0.41 (1.03) | -0.73 (0.49) | t = 10.19 | <.001 |

### 2.2. Study Design

Data were drawn from the Netherlands Study of Depression and Anxiety (NESDA), a multi-site naturalistic cohort examining the long-term course of depressive and anxiety disorders (3). Between 2005 and 2007, eligible participants were recruited into the NESDA Neuroimaging sub-study at three sites (Amsterdam, Leiden, Groningen (7)). Resting-state fMRI and diagnostic status were acquired at baseline (y0), two-year (y2), and nine-year (y9) follow-up.

### 2.3. Clinical measurements

At each session (y0, y2, y9), we also administered the Inventory of Depressive Symptomatology (IDS; (23)), Beck Anxiety Inventory (BAI; (24)), and Fear Questionnaire (25) to assess depression, anxiety, and phobia severity, respectively.

### 2.4 MRI image acquisition and processing

Data were acquired on 3T Philips MRI scanners at three centers in the Netherlands. Acquisition, preprocessing (fMRIPrep 24.0.1; (26), and denoising (XCP-D; (27)) protocols were identical across sites and timepoints. The full dataset was harmonized with the Longitudinal ComBat software (28). The implemented procedures are fully described in the “Supplementary Methods” section, SI.

### 2.5. Dynamic functional connectivity analysis

#### 2.5.1. Co-activation patterns (CAPs)

CAP analysis clusters individual resting-state fMRI volumes to identify recurring whole-brain transient co-(de)activation states (11,29). Unlike static functional connectivity, CAPs capture moment-to-moment spatial configurations represented as whole-brain z-scored maps. Brain CAPs were computed on data from all participants, pooled across the three sessions collected over nine years. See pipeline in Figure 2.

**Figure 2.**
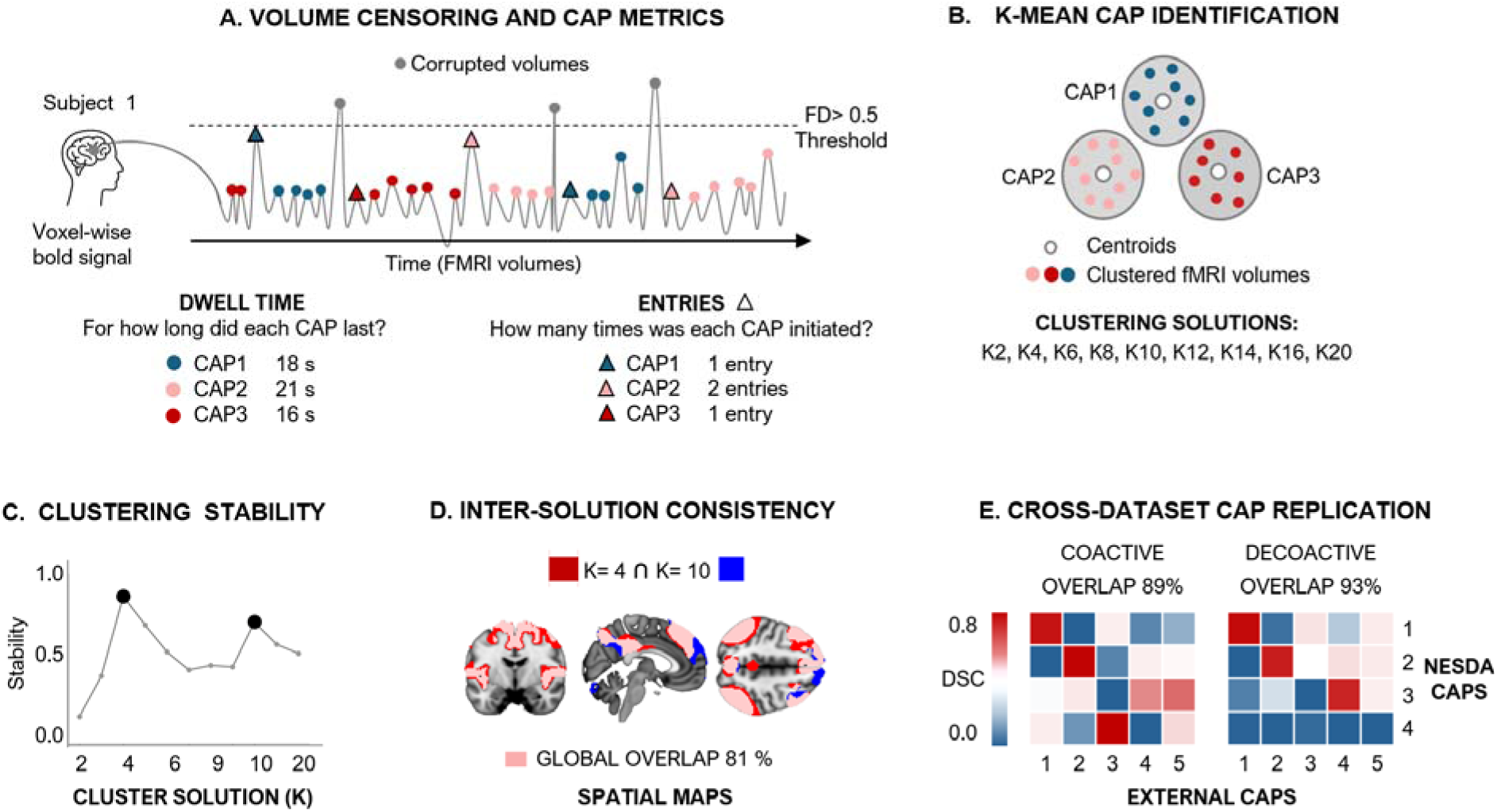
CAP extraction pipeline and solution validation. A. Volume censoring and CAP metrics. For each subject, framewise displacement (FD) was computed across the resting-state timeseries. Volumes with FD > 0.5 mm (corrupted volumes, gray dots) were excluded before clustering. Retained volumes were assigned to CAP states and characterized by two temporal metrics: dwell time (mean duration of consecutive frames in a given state, in seconds) and entries (number of times a state was initiated). **B. K-means CAP identification.** Retained volumes were subjected to K-means clustering across a range of K values (K = 3–12). Each solution partitions the fMRI volumes into K spatially distinct co-activation patterns defined by their centroids. **C. Clustering stability.** Stability was assessed across K using split-half resampling. K = 4 and K = 10 exhibited the highest stability coefficients (filled circles), motivating the selection of K = 4 as the primary solution due to noisy CAPs captured by K=10 (Figure S2A). **D. Inter-solution consistency.** Spatial overlap between the K = 4 and K = 10 solutions was examined. A global overlap of 81% across spatial maps confirms that the K = 4 states are reproduced within the finer-grained solution, supporting their robustness. **E. Cross-dataset CAP replication.** NESDA CAPs (rows 1–4) were spatially compared against an independent external CAP solution (columns 1–5) using the Dice Similarity Coefficient (DSC). Coactive (positive) and de-coactive (negative) spatial maps showed 89% and 93% overlap, respectively, confirming cross-dataset replicability of the K = 4 solution.

##### 2.5.1.1. CAPs characterization and validation

Optimal cluster number was determined via consensus k-means stability analysis ((30). Supplementary Methods S5.1). Internal stability was confirmed via spatial Dice overlap across solutions (S5.2; Figure S1). External validity was assessed against an independent dataset ((16); N = 116) via voxel-wise dice coefficients between matched CAP maps (S5.3; Figure S2). Network (i.e., CAP) composition was quantified as the proportion of co-activated (z > 1) and de-activated (z < −1) voxels overlapping canonical resting-state networks (Schaefer-400 + Harvard-Oxford subcortical atlas; 421 parcels; (31,32).

##### 2.5.1.2. Temporal metrics

To quantify subject-specific CAP expression over time, two temporal metrics were computed: “entries” (counts; number of times a person transitioned into a given CAP (33), indicating how many times a CAP was initiated (Figure 2A); and “dwell time” (continuous, in seconds; see “persistence” in (17) or “duration” in (18)), the average duration of each CAP episode, i.e., the average time spent in a CAP once entered.

### 2.6 Statistical Analysis

#### 2.6.1. Baseline cross-sectional analysis

To test our first hypothesis (H1), the expression of brain CAPs (dwell time [DUR], entries [ENT]) and symptom severity (depression [IDS], anxiety [BAI], and fear [FQ]) were first assessed respectively. Mann–Whitney U tests, with rank-biserial correlation reported as the effect size, were applied to z-scored variables to assess difference between the AFF (*N* = 228) and CTR (*N* = 112) group in terms of these variables. Baseline CAP–symptom associations were then quantified with partial Spearman correlations, computed in the full pooled sample (*N* = 317). All *p*-values were corrected for false discovery rate (BH-FDR) across 4 CAPs × 3 symptoms within each group. Correlations were adjusted for age, sex, drug use, scanning site, education level, and group (AFF, CTR).

#### 2.6.2 Longitudinal analysis

As a preliminary step, group (AFF, CTR) × session (Y0, Y2, Y9) linear mixed models (LMMs) with a random intercept per subject tested CAP and symptom trajectories across the follow-up. The models were adjusted for sex, age, education level, and scanning site. Models Diagnostics verified the appropriateness of a combined model across groups: Levene’s test on residuals and a group-specific residual-variance model (compared by AIC and likelihood-ratio test) assessed the equal-variance assumption, while singular-fit checks and per-group residual-normality tests confirmed the random-effects structure was stable (Supplementary Methods S5). All models were implemented in R, lme4/lmerTest (REML, Satterthwaite df).

##### CAP–symptom coupling

To test our second hypothesis (H2), whether CAP dynamics (dwell time [DUR], entries [ENT]) track the longitudinal course of depression (IDS), anxiety (BAI), and fear (FQ), we applied three complementary within-person analyses. A first set of LMMs (“symptoms x group (AFF, CTR)” interaction) Assessed whether within-person coupling (i.e., the link between a brain-state (CAP) and symptom fluctuations, estimated inside each individual) differs between AFF and CTR (group-specificity). A second set of LMMs (“symptoms x session (Y0, Y2, Y9)” interaction), examined CAP-symptom coupling referenced to each person’s mean symptom level (i.e., whether, at sessions where a subject’s symptoms are above or below their personal average, CAP expression is correspondingly above or below average (time-stability)). Third, change–change Spearman correlations estimated coupling in the direction of change between sessions. Namely, whether changes in CAP were concurrent with changes in symptom severity (directional, interval-specific). All the models were adjusted for sex, age, drug use, education level, and scanning site.

##### Linear mixed modelling (LMM)

For each symptom score, we decomposed the repeated measures into a between-person component (each subject’s average across their available sessions) and a within-person component (each session’s deviation from that mean). Both were entered as predictors of the rank-transformed CAP metrics (dwell time, entries) in the LMMs with a random intercept per subject; ranking the CAP outcome provides a nonparametric estimate robust to its bounded, non-normal distribution. Group-specificity and time-stability were tested by “symptom×group” and “symptom×session” interaction terms, respectively (as defined above). Satterthwaite degrees of freedom were used, and effect sizes were partial eta-squared (η²p). Interaction p-values were FDR-corrected (Benjamini–Hochberg) across each analysis’s full grid of CAP × symptom tests (4 CAPs × 3 symptoms). Full model specification, the within-between decomposition, and fit diagnostics are described in Supplementary Methods S5.

##### Change–change correlations (rho)

Each subject contributed one paired difference per interval. Namely, changes in CAP (dwell time, entries), against the change in symptoms (IDS, BAI, FQ) from one timepoint to the next (Y0→Y2, Y0→Y9, Y2→Y9)). These interval-specific correlations were computed separately in the AFF and CTR groups. Here the reference is the subject’s previous timepoint. These Spearman rho models examined whether, when a person moved from one session to the next, CAP and symptoms shifted in the same direction. Multiple comparisons were controlled with Benjamini–Hochberg FDR within each group × symptom family (4 CAPs × 3 symptoms × 3 intervals).

## 3. RESULTS

### 3.1 Sample demographics

The analytic sample comprised 340 participants with usable resting-state fMRI at one or more sessions: 228 with a current affective disorder (AFF; MDD and/or an anxiety disorder) and 112 controls (CTR). The AFF group was modestly younger than CTR (38.7 vs. 42.5 years) and less often university-educated, while sex distribution was comparable; the two groups differed markedly on all three symptom dimensions (all *p* < 0.001; Table 1). Because neuroimaging was acquired on a rolling basis, each participant’s baseline was defined as their earliest available session rather than a fixed calendar timepoint, so a given assessment wave (Year 0, Year 2, Year 9) combines participants entering imaging for the first time with those returning for follow-up (Figure 1).

### 3.2. Identification and validation of co-activation patterns

K-means clustering was applied to the full fMRI dataset across solutions K=2–20, evaluated via consensus split-half stability resampling. K=4 and K=10 yielded the highest stability coefficients (Figure 2C). K=4 was selected as the primary solution because K=10 introduced two spatially noisy, uninterpretable states (Figure S2A), whereas each K=4 CAP was internally replicated within the K=10 solution (mean 81% spatial overlap, Dice; Figures S2A, 1D). This shows that the four coarser states are not arbitrary groupings but reproducible patterns that persist even when the data are partitioned more finely. External cross-validation against an independent dataset (15) demonstrated high spatial replicability of both co-active and de-active spatial maps (mean Dice 89% and 93%, respectively; Figure 2E). Network composition analysis (Schaefer-400 + Harvard-Oxford atlas, 421 parcels) yielded four spatially distinct states, labelled AT-SM_CAP_ (CAP1), DM_CAP_ (CAP2), FP-DM_CAP_ (CAP3), and VS-SC_CAP_ (CAP4) based on their dominant network overlap (Figure 2A).

### 3.3. Spatial representation of brain CAPs

CAP1 showed co-activation of sensorimotor, salience/ventral attention, temporoparietal, and frontoparietal control regions, with sensorimotor and salience accounting for 65% of co-activation; we labelled this state Attention–Sensorimotor CAP (**AT-SM_CAP_**; Figure 3A; Table S1). Co-deactivation primarily involved default mode regions, including medial prefrontal cortex, precuneus/PCC, and bilateral angular gyri (Table S1). CAP2 (**DM_CAP_**) reflected co-activation of default mode subsystems including medial prefrontal, precuneus/PCC, angular gyri, hippocampus, and lateral temporal cortex, alongside limbic, cerebellar, and subcortical regions, with DMN accounting for 60% of co-activation. De-coactivation encompassed dorsal and ventral attention, lateral frontoparietal control, and sensorimotor regions (Figure 3A; Table S2). CAP3 (**FP-DM_CAP_**) showed co-activation of frontoparietal control (49%) and default mode (25%) regions, including bilateral lateral prefrontal cortex, inferior parietal lobule, angular gyri, precuneus, and mid-cingulate. Co-deactivation involved the entire visual cortex and bilateral sensorimotor strip (Figure 3A; Table S3). CAP4 (**VS-SC_CAP_**) comprised co-activation of visual, dorsal attention, somatomotor, and subcortical/cerebellar regions spanning the posterior cortex bilaterally, with no co-deactivation clusters surviving threshold (Figure 3A; Table S4). All four CAPs were internally reproduced within the K=10 solution and externally replicated in an independent dataset (16), confirming their robustness and reproducibility (full validation statistics in Supplementary Methods S4, Figure S2).

**Figure 3.**
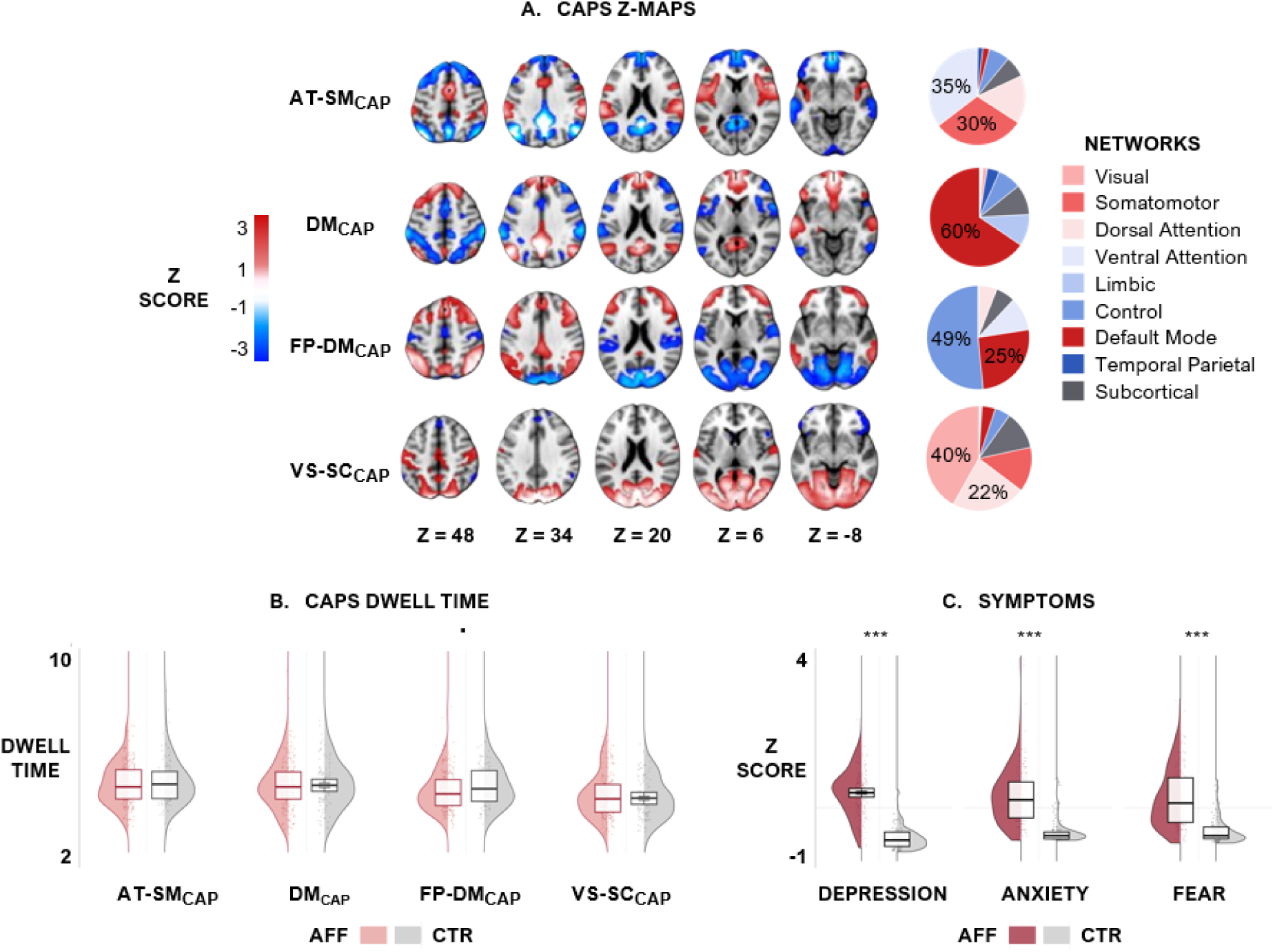
Spatial maps, dwell time distributions, and baseline symptom profiles of K=4 co-activation patterns. **(A) CAP Z-maps.** Group-level z-score activation maps for each of the four CAPs (rows) across five axial slices (Z = 48, 34, 20, 6, −8). Warm colors (red) indicate positive z-scores; cool colors (blue) indicate negative z-scores. Pie charts (right) show the proportional contribution of canonical resting-state networks to each CAP’s positive activation. CAP labels reflect dominant network composition: AT-SM (Attention–Somatomotor), DM (Default Mode), FP-DM (Frontoparietal–Default Mode), VS-SC (Visual–Subcortical). **B. CAP dwell time at baseline.** Violin plots with embedded box plots showing the distribution of mean dwell time (seconds) for each CAP separately for affective disorder patients (AFF; pink) and healthy controls (CTR; gray) at baseline (W1). The dot between FP-DM_CAP_ violin plots indicates a trend-level group difference. Group differences in CAP entries were nonsignificant. **C Baseline symptom severity.** Violin plots with embedded box plots showing z-scored depression, anxiety, and fear scores at baseline for AFF (dark red) and CTR (gray). Asterisks indicate significant group differences (*** *q* < .001).

### 3.4. Cross-sectional baseline analysis

As preliminary assessment, Mann–Whitney U tests compared AFF and CTR on CAP dwell duration, entries, and symptom severity individually at baseline (Table S5). Dwell time and entries were each similar within AFF and within CTR across the four CAPs (AFF: ∼4.2–4.8 s dwell, ∼24–26 entries; CTR: comparable ranges). FP-DM_CAP_ dwell time showed the closest approach to a group difference (trend-level, *q* = 0.09; Fig. 3B). Symptom severity differed markedly between groups on all three measures (all *q* < .001; Fig. 3C).

#### 3.4.1. Baseline coupling between CAP dynamics and symptoms severity

To test our first hypothesis (H1), that brain temporal dynamics are associated with symptom severity cross-sectionally, we computed Spearman correlations between CAPs (dwell time [DUR], entries [ENT]) and symptom severity (depression [IDS], anxiety [BAI], and fear [FQ]) in the pooled sample (*N*= 317). Group membership had the largest effect of any covariate on the estimates, and the pooled-data associations proved spurious due to Simpson’s paradox ((34) Fig. S3). Group-stratified analysis revealed AT-SM_CAP_ dwell time was negatively associated with depression severity in CTR (N = 92, rho = −0.29, 95% CI [−0.47, −0.08], *q* < 0.05), but not in AFF (N = 225, rho = 0.09, *q* = 0.79; Fig. 4B; full results in Table S6A). Sensitivity analyses confirmed both results: in CTR, leave-one-out estimates (LOO) ranged from −0.31 to −0.26 (no single outlier drove the effect), while trimming the upper symptom range attenuated rho smoothly (top 5%: −0.24; top 10%: −0.18; top 15%: −0.13), consistent with reduced power in a shrinking, more homogeneous subsample. In AFF, rho remained stable regardless of trimming direction or extent (LOO results: 0.05–0.11 across all checks), and its bootstrap 95% CI spanned zero [−0.04, 0.22], confirming the null was not masking a true effect. Associations between CAP entries and symptoms were nonsignificant (Table S6B). **Group differences in CAP–symptom coupling at baseline.** Because inferring group differences from a significance threshold is a statistical fallacy (e.g., one correlation significant (*p*<.05) but not the other (*p*> 0.05) (35)), Fisher z-tests directly tested differences in CAP–symptom coefficients (Δ = rhoAFF − rhoCTR). The AT-SM_CAP_–IDS group contrast was significant (Δrho = 0.36, Z = 2.99, 95% CI [0.13, 0.56], *q* < 0.05). All remaining contrasts were non-significant (*q* > 0.05 Fig. 3C, 3D). Covariate adjustment had minimal impact.

**Figure 4.**
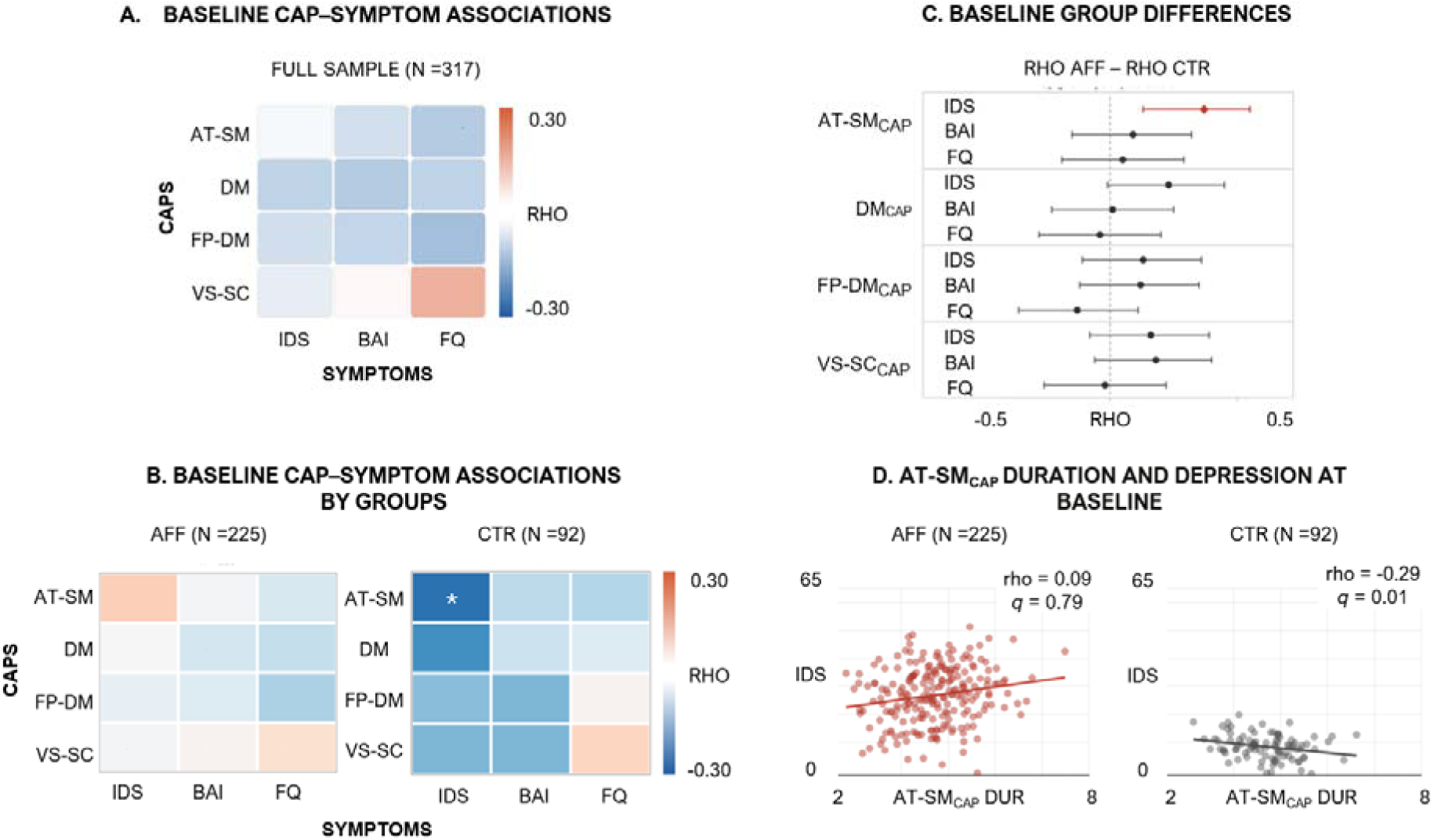
Cross-sectional analysis. Associations between CAP dwell duration and symptom severity at baseline. **A.** Spearman correlations (rho) between dwell duration of each brain CAP and three symptom dimensions—depression (IDS), anxiety (BAI), and fear (FQ)—in the full baseline sample (N = 317). **B.** Correlations stratified by affective patients (AFF, n = 225; left) and controls (CTR, n = 92; right). The asterisk indicates the strongest association overall (AT-SM_CAP_–IDS in CTR: rho = −0.29, *q*< 0.05). **C.** Group differences in CAP–symptom associations (Δ = rho_AFF_ − rho_CTR_) assessed with Fisher z-tests (N_AFF_ = 225, N_CTR_ = 92; SE = 0.125), plotted with 95% confidence intervals on Δ; the dashed line indicates zero. Correlations between AT-SM_CAP_ and IDS reported the greatest difference in their coefficients (*q* < 0.01). **D.** Scatterplots illustrate the AT-SM_CAP_ dwell duration × depression association in AFF (rho = 0.09, *q* = 0.79) and CTR (rho = −0.29, *q*< 0.05), with regression lines for visualization. *q*: FDR-corrected *p*-values across symptom x CAP associations and group comparisons reported in the figure.

### 3.5. Longitudinal analysis

Here, we examined our second hypothesis (H2), whether within-person fluctuations of CAP dwell time and entries track concurrent symptom severity changes over nine years.

#### 3.5.1. Nine-year trajectories

Group × session interactions tested whether symptoms and CAPs trajectories differed in shape between AFF and CTR. **Symptoms:** depression and anxiety trajectories differed significantly by group (IDS: *F*=7.84, *df*=2,39, *q* = 0.001, η*²*p=.04; BAI: *F*=5.80, *df*=2,359, *q*=.005, η*²p*=0.03. table S7A, S7B), driven by improvement in AFF from Y0 to Y2 (*d* = −0.46, −0.34; Table S8) while CTR remained low and stable (Table S9); fear trajectories did not differ by group (*q* = 0.22. Table S7). **CAPs:** no CAP metric showed a significant group × time interaction, for dwell time or entries, in any of the four CAPs (all *q* ≥ 0.83; Table S7A, S7B). CAP dwell time showed similar wave-to-wave fluctuation in both groups — e.g., FP-DM_CAP_ rebounded between Y2 and Y9 in AFF (*d* = 0.57–0.88; Table S8) and equally in CTR (*d* = 0.66; Table S9), consistent with the absence of a group-specific trajectory shape.

#### 3.5.2. Longitudinal coupling between CAP dynamics and symptoms severity

Diagnostics supported the pooled group-interaction model specification (Tables S10a, S10b). In the combined within-between LMMs, within-person coupling did not differ between AFF and CTR for any CAP–symptom pair (symptom-by-group interaction, all *q* ≥ 0.54, all η²p ≤ 0.008; Table S11), and the session-moderated interaction was likewise null after correction in both groups (all *q* ≥ 0.27; Table S12A, S12B). Because these models reference coupling to each person’s mean symptom level and can miss coupling expressed as directional change between sessions, we examined interval-specific change–change correlations (rho(ΔCAP, Δsymptom); Y0→Y2, Y0→Y9, Y2→Y9), in AFF and CTR respectively.

##### ΔFP-DM_CAP_ dwell time and ΔIDS

This coupling emerged exclusively in the AFF group, over y2→y9 (*N* = 39), FP-DM_CAP_ dwell time increased on average (Δz = 0.62; *p* = 0.011) while depression was unchanged at the group level (Δz = −0.05; *p* = 0.61); within persons, however, changes in the two measures co-varied positively (rho = 0.47, 95% CI [0.18, 0.68], *q* < 0.01; unadjusted rho = 0.50, *p* = 0.001; Fig. 4B. Table S13). This effect was absent over y0→y2 (rho = 0.02, 95% CI [−0.19, 0.24]) and y0→y9 (rho = −0.05, 95% CI [−0.33, 0.24]). Cross-sectional coupling was null at both Y2 and Y9 (|rho| ≤ 0.13), consistent with a within-person rather than between-person effect. The Y2→Y9 coupling was similarly absent in CTR (rho = 0.11, *q* > 0.05). This null result was stable under sensitivity checks (leave-one-out rho 0.01–0.24; bootstrap 95% CI [−0.31, 0.51]), consistent with limited power (*N* = 21) rather than a masked effect. A sensitivity analysis restricted to participants with data at all three sessions confirmed the finding (*N* = 32; Fig. S4A).

##### ΔVS-SC_CAP_ dwell time and ΔFQ

A positive association between VS-SC_CAP_ dwell time change and fear change emerged, again, only in the AFF group over y0→y9 (N_AFF = 49); absent in CTR: rho = 0.15, *q* > 0.05, stable under sensitivity checks — leave-one-out rho 0.04–0.27, bootstrap 95% CI [−0.39, 0.60]), absent over y0→y2 (rho = 0.08, 95% CI [−0.29, 0.14]) and y2→y9 (rho = −0.14, 95% CI [−0.44, 0.18]. This association was not confirmed in the sensitivity analysis restricted to participants with complete three-wave data: the effect attenuated to non-significance (rho = 0.24, N_AFF_ = 32; Fig. S4B), partly attributable to participants who attended y0 and y9 but not y2 and showed disproportionate clinical improvement over the full interval. Although the directionality was preserved, this finding should be interpreted with caution. Across all remaining CAP × symptom × interval combinations, one additional association reached nominal significance: DM_CAP_ entries and fear over y0→y9 only in the AFF group, but did not survive FDR correction across the 36 tests conducted and is not interpreted further.

## 4. DISCUSSION

We used a data-driven co-activation pattern (CAP) approach to test whether whole-brain dynamic states relate to affective symptom severity, both cross-sectionally and across nine years of naturalistic follow-up. Four reproducible CAPs were identified, internally validated and externally replicated. Two findings emerged. Cross-sectionally, longer AT-SM_CAP_ dwell time was associated with lower depression in controls but not in patients (H1). Longitudinally, within-person CAP dwell time fluctuations tracked the failure to improve in depression and fear, exclusively in affected individuals (H2). Together, these findings link the persistence of whole-brain states to affective symptoms at two complementary levels: A cross-sectional marker of health versus illness, and a within-person index of illness course.

### 4.1. Cross-sectional CAP–symptom associations (H1)

At baseline, CAP metrics did not differ between groups, and pooled-sample correlations were uniformly small. One robust association emerged only after stratification: in controls, greater AT-SM_CAP_ dwell time was negatively associated with depression severity, an effect absent in patients and yielding a significant between-group difference in coupling (Δ = rhoAFF − rhoCTR). Because AT-SM_CAP_ co-activates sensorimotor, salience, and attentional control regions while de-activating the default mode (36), prolonged occupancy of this task-positive state may index a mood-regulatory capacity (15,37) operative in health but uncoupled in affective disorders (15,38). Whereas controls sustain this state in proportion to lower symptom burden, patients show no such inverse relationship, consistent with a loss of adaptive affective regulation (39).

### 4.2. Longitudinal within-person coupling (H2)

Two rank-based linear mixed models (§3.5.2) tested whether within-person CAP-symptom coupling differed between AFF and CTR, and whether it varied across sessions. Their results yielded weak associations. However, interval-specific correlations between changes in CAPs expression (ΔCAPs) and changes in symptoms severity (Δsymptoms) revealed a robust within-person coupling in the AFF group: FP-DM_CAP_ dwell time tracked worsening depression from Year 2 to Year 9. Notably, these methods pursued different aims. Both LMMs quantified within-person coupling referenced to each person’s own mean symptom level (i.e., whether sessions with below/above average symptoms also showed below/above average CAP expression). One testing whether that relationship differed by group. The other, whether the CAP-symptom coupling differed by session. By contrast, the change–change coupling (rho(ΔCAPs, Δsymptoms)) asked whether, within each patient, a shift in the expression of CAPs tracks a shift in symptoms severity over the same interval, capturing within-person co-variation across time.

#### ΔFP-DM_CAP_ – Δdepression coupling

FP-DM_CAP_ regions have each been linked to depression and rumination: the dorsolateral prefrontal cortex, whose activity scales with symptom severity (40); angular and inferior parietal cortices, implicated in rumination (41); and cingulate–precuneus default-mode hubs central to self-referential processing (42). Its de-coactivation map spans the visual–somatomotor territory that constitutes the VS-SC_CAP_ coactivation state. To our knowledge, this is the first report of FP-DM dynamic connectivity in affective disorders. The reports mentioned linking this system to depression relied on stationary connectivity. This result suggests that longer-sustained occupancy of this state accompanies failure to remit, consistent with an inability to disengage from perseverative self-referential processing as illness continues. The ΔFP-DM_CAP_ – Δdepression coupling was confirmed and numerically strengthened in sensitivity analyses restricted to participants with fMRI and clinical data in the three sessions.

Why did changes in FP-DM_CAP_ correlate with depression changes only at Y2→Y9? Its emergence specifically over this interval highlights the importance of within-person approaches in the assessment of brain–symptom interactions. At y0→y2, despite the largest symptom improvement, individuals showed no consistent FP-DM_CAP_ change, indicating early recovery occurred independently of this brain state, and likely reflecting the heterogeneity of recovery pathways in the naturalistic NESDA sample. By y2→y9, when early symptom changes had stabilized, 69% of individuals showed concordant ΔFP-DM_CAP_/ΔIDS directionality (versus 53% at y0→y2), and those whose dwell time rebounded were precisely those whose depression failed to improve further. The Y0→Y9 interval attenuated this coupling because FP-DM_CAP_ changes over y0→y2 and y2→y9 moved in opposing directions within the same individuals. Group-level trajectories did not capture this. Symptoms merely plateaued at the group mean (Figure 5A), underscoring that the coupling is a within-person phenomenon invisible to population-averaged observations.

**Figure 5.**
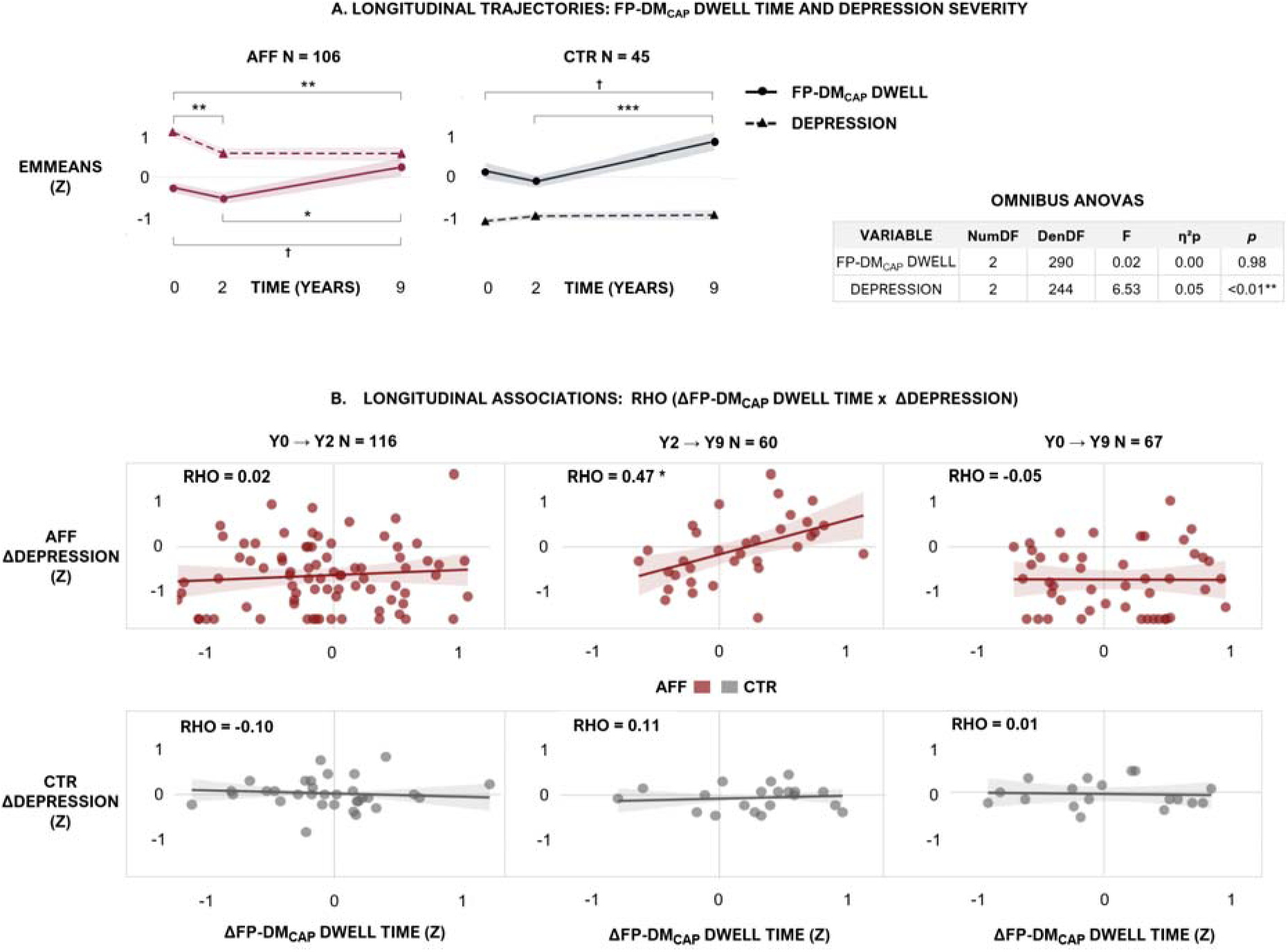
Longitudinal associations between CAP dwell duration and symptom severity over 9 years. A. Group-mean trajectories of FP-DM_CAP_ dwell time and depression severity. Estimated marginal means (±1 SE; z-scored) of FP-DM_CAP_ dwell time (solid line, circles) and depression severity (dashed line, triangles) at baseline (y0), 2-year follow-up (y2), and 9-year follow-up (y9), in affective patients (AFF, dark red; n = 106) and healthy controls (CTR, grey; n = 45). Omnibus repeated-measures ANOVAs confirmed that depression showed significant within-person variability across the three sessions (F(2, 244) = 6.53, η²p = 0.05, p < 0.01), whereas FP-DM_CAP_ dwell time did not (F(2, 290) = 0.02, η²p < 0.01, p = 0.98). In AFF, depression improves markedly over 9 years, while FP-DM_CAP_ dwell time rebounds between y2 and y9. Significance markers denote pairwise interval contrasts from the within-group LMM, FDR-corrected across the three-time contrasts (y0→y2, y2→y9, y0→y9) per variable and group (AFF, CTR). **B. Interval-specific change–change coupling.** rho(ΔFP-DM_CAP__DUR × ΔDEPRESSION). Each point represents one subject. X-axis: within-person change in FP-DM_CAP_ dwell time (Δz); y-axis: within-person change in depression severity (Δz) over the same interval. Top row: AFF; bottom row: CTR. Columns correspond to three intervals: y0→y2 (early recovery), y2→y9 (late follow-up), and y0→y9 (full 9-year span). The coupling emerges exclusively at y2→y9, absent at y0→y2 and y0→y9, and exclusively in AFF. A pattern not predicted by the group-mean trajectories in Panel A. The regression line is derived from the fully adjusted Spearman rho(covariates: age, sex, drug use, scanning site, education level). Shaded band: 95% confidence interval of the regression line (Fisher Z-transformation). Reported rho reflects the complete interval sample. AFF: y0→y2 n = 82, y2→y9 n = 39, y0→y9 n = 49; CTR: y0→y2 n = 34, y2→y9 n = 21, y0→y9 n = 18. In Panel A, † q < 0.10, * q < 0.05, ** *q* < 0.01, *** *q* < 0.001; FDR-corrected across the three pairwise time contrasts (y0→y2, y2→y9, y0→y9) within each variable and group independently. In Panel B, ** *q* < 0.01, * *q* < 0.05; FDR-corrected across all CAP × symptom × interval combinations (36 tests) within each group independently.

Further associations were observed across the full nine-year interval (y0→y9). The coupling between VS-SC_CAP_ dwell time and persistent fear was directionally consistent with ΔFP-DM_CAP_ – Δdepression coupling, but less robust and sensitive to sample composition: when restricted to participants with complete three-session fMRI data, the effect attenuated to non-significance, partly attributable to participants who attended y0 and y9 but not y2 and showed disproportionate clinical improvement over the full interval. Additionally, changes in DM_CAP_ entries also tracked fear over the same interval, but this coupling did not survive correction for multiple testing. Notably, this DM_CAP_ entries – fear coupling has been independently linked to fear reduction under running therapy (16).

## 5. LIMITATIONS

Several constraints temper our interpretation. The longitudinal couplings rested on modest interval samples. However, longitudinal attrition is expected in 9-year psychiatric studies. Within-person change analyses (N=67 for full span) retain adequate statistical power to detect the observed coupling effects while capturing long-term illness trajectories. CAP robustness (internal and external testing) ensured findings reflect neurobiology, not measurement artifact. Effect sizes for the cross-sectional association were modest and warrant replication in independent cohorts. The change–change framework, although well suited to isolating within-person co-variation, cannot establish the temporal precedence of brain change relative to symptom change, leaving the directionality of the coupling open. Finally, the naturalistic design captures illness course under heterogeneous, uncontrolled treatment, which both grounds the findings in real-world trajectories and limits mechanistic inference. Also, The MRI subsample size limited our ability to examine affective disorder types (e.g., depression and anxiety).

## 6. CONCLUSIONS

Across nine years of naturalistic follow-up, CAP dwell-time–symptom coupling showed clinical relevance at two levels: as a between-subject marker that distinguishes health from illness cross-sectionally, and as a within-person index of illness course longitudinally that is not apparent in group-level or between-person analyses. Dwell time in the brain frontoparietal–default-mode system (FP–DM_CAP_) tracked worsening depression exclusively within individuals suffering from affective disorders (AFF). While self-report instruments reliably capture symptom course, CAP–symptom coupling adds a neurobiological dimension by identifying which individuals’ brain dynamics mirror their illness trajectories. By grounding symptom change in a reproducible whole-brain signature at the individual level, this coupling provides a candidate biomarker to refine diagnosis and guide individually targeted interventions, consistent with the within-person focus of precision psychiatry (21).

## 7. ACKNOWLEDGMENTS

Special thanks to Thomas Bolton for the original CAPs toolbox code. An adapted version is available at https://github.com/JulianGaviriaL/NESDA.

## 8. DATA AVAILABILITY

Data are available through formal requests (analysis plan) submitted to: https://www.nesda.nl/researchers/nesda-pro/data-and-documentation/. Analysis code is available at https://github.com/JulianGaviriaL/NESDA.

## 9. DECLARATION OF AI-ASSISTED TECHNOLOGIES

The authors used Claude (Sonnet 4.6) for manuscript preparation and take full responsibility for the final content.

