## Supplementary material for "Brain functional dynamics linked to depression and anxiety: cross-sectional and longitudinal associations over 9 years"

Gaviria Lopez *et al.*

### **Supplementary methods**

#### **S1. fMRI Preprocessing**

**S1.1 Structural MRI Processing**

Structural MRI images were skull-stripped and segmented to reconstruct brain surfaces using FreeSurfer 7.3.2. For subjects with longitudinal data, we generated a robust mean template of all available T1-weighted (T1w) scans using *mri_robust_template*; otherwise, the T1w image at baseline was used. Bias field correction was applied using N4BiasFieldCorrection (ANTs 2.5.1), and brain tissue segmentation (CSF, white matter, grey matter) was performed with FAST (FSL). Spatial normalization to MNI152NLin6Asym and MNI152NLin2009cAsym standard spaces was performed via nonlinear registration using antsRegistration (ANTs 2.5.1), with transformations derived using TemplateFlow (1).

**S1.2 Functional MRI Preprocessing.**

Resting-state fMRI images were preprocessed using fMRIPrep 24.0.1 (2). Briefly, BOLD images were skull-stripped, realigned (6 degrees-of-freedom rigid-body head motion estimation using *mcflirt*, FSL), and corrected for susceptibility-induced distortions using a fieldmap-less approach (SyN: EPI-to-T1w co-registration with intensity inversion and phase-encoding–constrained deformation). BOLD images were co-registered to the subject's T1w image using boundary-based registration (*bbregister*, FreeSurfer), and resampled in a single interpolation step to MNI152NLin6Asym space at 2 mm isotropic resolution. Global signal regression (GSR) was not applied, given the long-standing disagreement on this procedure in the field. While some studies support its use, GSR has been shown to introduce spurious anticorrelations and to distort between-group covariance structure — a particular concern for longitudinal between-treatment contrasts. Importantly, prior work has shown that GSR does not meaningfully affect the CAP extraction procedure, and that CAP-based estimates of network anticorrelation remain stable regardless GSR implementation (3). We therefore relied on the benchmarked aCompCor + cosine denoising strategy described below. **Quality Control**

Image quality metrics (DVARS and temporal SNR) were calculated using MRIqc and compared between groups and timepoints to ensure no systematic quality differences.

#### **S2. Post-Processing denoising.**

The eXtensible Connectivity Pipeline-DCAN (XCP-D version 0.14.1; (4) was used to post-process the outputs of fMRIPrep 24.0.1. XCP-D was built with Nipype version 1.10.0. **Input:** fMRIPrep BOLD derivatives (NIfTI format, MNI152NLin6Asym space, 2 mm isotropic). Each BOLD run was processed independently. Native-space T1w images were transformed to MNI152NLin6Asym space at 1 mm³ resolution using the nonlinear warp from fMRIPrep.

**S2.1. Denoising Pipeline.** The following sequential denoising steps were applied to each BOLD run:

**Step1. Despiking.** Large intensity transients were removed using AFNI's *3dDespike* prior to regressions.

**Step2. Interpolation.** Censored frames were temporarily replaced via cubic spline interpolation. Edge volumes were replaced with the nearest low-motion frame's values to prevent extrapolation artefacts.

**Step3. Nuisance Regression (aCompCor strategy).** A single GLM was estimated using the following regressors: (i) top 5 aCompCor principal components from the white matter compartment; (ii) top 5 aCompCor principal components from the CSF compartment; (iii) 6 rigid-body motion parameters and their temporal derivatives (12 total); (iv) cosine basis regressors (session-length dependent), acting as a high-pass filter via regression rather than frequency-domain filtering. The GLM was estimated on low-motion volumes only and applied to the full interpolated time series. No bandpass filter was applied; the cosine regression replaces frequency-domain filtering. Global signal was explicitly excluded from the regressor set for avoiding spurious brain regions interactions in further steps (5).

**Step4. Censoring.** FD was calculated using the Power et al. formula (6), with head radius auto-estimated per subject. Volumes with FD > 0.5 mm were flagged as high-motion outliers for subsequent censoring. No motion filtering was applied. A minimum temporal coverage criterion required that at least 85% of frames survive censoring.

**Step5. Spatial Smoothing.** The denoised, censored BOLD signal was spatially smoothed using a Gaussian kernel (FWHM = 6 mm; Nilearn 0.13.0).

**S2.4 Additional Quality Metrics.** Regional Homogeneity (7) was computed with neighborhood voxels using AFNI's 3dReHo (8). Many internal operations of XCP-D use AFNI, ANTs, TemplateFlow, matplotlib, Nibabel, Nilearn, NumPy, PyBIDS, and SciPy.

**S3. Denoising Pipeline Validation.** To verify that the XCP-D denoising pipeline performed as intended, three quantitative validation tests were applied to one participant randomly selected comparing the fMRIPrep-preprocessed BOLD signal (before denoising) to the XCP-D denoised output (after denoising).

**S3.1 Test 1: Power Spectrum Comparison (Before vs. After Denoising.** **Figure S1A**)**.** To verify that denoising removed the correct frequency components without destroying neural signal, we extracted the mean whole-brain BOLD timeseries before and after denoising and computed the Fast Fourier Transform (FFT) power spectrum for each. Before denoising, the spectrum shows high broadband power characteristic of mixed signal and noise. After denoising with the aCompCor + cosine strategy, three expected changes are observed: (i) the absence of the large low-frequency power spike that dominates the before-denoising spectrum (reaching 10¹⁰ near 0 Hz; Figure S1A, left). After denoising Figure S1A, right), power does not surge at near-zero frequencies but instead rises gradually to a stable plateau by ~0.01 Hz, confirming that scanner drift — the dominant source of low-frequency contamination — was effectively removed by cosine regressors; (ii) a flat plateau from 0.01–0.20 Hz, confirming that neural signal in the BOLD frequency band is preserved; and (iii) an overall reduction in spectral power without selective attenuation of the physiologically relevant band, indicating targeted rather than indiscriminate noise removal.

**S3.2 Test 2: Motion–Signal Coupling (Figure S1B).** To confirm that head motion artefacts were decoupled from the BOLD signal, we computed framewise displacement (FD) from the fMRIPrep confound file and framewise signal change (DVARS) before and after denoising, then quantified their Pearson correlation. Before denoising, FD–DVARS correlation was r = 0.053 (variance explained: 0.28%). After denoising, the correlation dropped to r = 0.003 (variance explained: 0.00%), a 94% reduction. Motion-explained variance was effectively eliminated, confirming that head movement no longer drives BOLD signal fluctuations in the denoised data.

**S3.3 Test 3: Temporal Variance Reduction (Figure S1C)** To assess the magnitude of noise removal without over-regression, we computed the temporal standard deviation (tSD) of every brain voxel before and after denoising and averaged within the brain mask. A 70–95% reduction was used as the pass criterion, with >98% flagged as potential over-regression. Mean tSD was reduced from 13,074 to 1,938 (85% reduction), and spatial variability of the tSD map decreased from 15,488 to 1,027 (93% reduction). Both values fall within the expected 70–95% range, confirming effective noise removal without evidence of over-regression. The remaining 15% of temporal variance constitutes the neural signal retained for CAP analysis.

**S3.4. Denoising Regressor Verification (Design Matrix).**

To confirm that XCP-D used the correct regressors in the denoising GLM, we inspected the design matrix output for the same representative participant. The expected regressor set for the aCompCor strategy comprises: 12 motion parameters (6 rigid-body parameters + 6 temporal derivatives), 5 WM aCompCor components, 5 CSF aCompCor components, and session-length–dependent cosine drift regressors. Global signal should be absent.

| **Regressor category** | **Expected count** | **Observed count** |
| --- | --- | --- |
| Motion parameters (trans/rot + derivatives) | 12 | 12 |
| aCompCor WM (w_comp_cor) | 5 | 5 |
| aCompCor CSF (c_comp_cor) | 5 | 5 |
| Cosine drift regressors | ~6 (varies) | Session-dependent |

**Design matrix verification for the representative participant (sub-110087, ses-ams03).** All expected regressor categories were present at the correct count, and global signal was absent, confirming correct implementation of the aCompCor denoising strategy.

**S4. Site Harmonization of CAP Temporal Metrics**

**S4.1 Rationale.** Although acquisition, preprocessing, and denoising protocols were identical across the three recruitment sites (Amsterdam, Leiden, Groningen; Sections S1–S3), a real hardware asymmetry exists across sites (SENSE 8-channel receiver coils at Leiden and Groningen vs. SENSE 6-channel at Amsterdam; main text Section 2.4), which could in principle introduce residual scanner-driven variance in the derived CAP temporal metrics. To test for and remove any such effect, we applied Longitudinal ComBat (9) to the two CAP temporal metrics, dwell time (DUR) and entries (ENT), computed for each of the four CAPs.

**S4.2 Model.** Harmonization was applied to the derived per-subject, per-wave CAP metrics, not to the raw BOLD signal or the CAP-defining clustering step. This follows the standard point of application for ComBat-family harmonization in the imaging literature: region- or feature-level harmonization downstream of a shared, sample-wide feature-extraction pipeline. CAP cluster centroids were estimated once across the pooled sample (Section S5.1), so the measurement instrument itself was identical across sites; harmonization instead targeted subject-level occupancy summaries computed under that shared instrument. For each metric (DUR, ENT) and each CAP, longitudinal ComBat was fit with site (3 levels: Amsterdam, Leiden, Groningen) as the batch variable, wave (Y0/Y2/Y9) as the repeated-measures time variable, a random intercept per subject to accommodate the unbalanced longitudinal design, and group, age, and sex protected as covariates (variance associated with these terms was not removed). Analyses used the longCombat R package (9).

**S4.3 Site-associated variance before and after harmonization.** Before harmonization, site explained a small proportion of variance across all eight CAP-metric combinations (R² range: <0.001–0.041; Table S15), consistent with the identical cross-site acquisition and denoising protocol described in Sections S1–S2. The largest raw effect was observed for VS-SC_CAP_ dwell time (R² = 0.041). After longitudinal ComBat harmonization, site-associated variance dropped to near zero across all combinations (R² ≤ 0.001; Table below), confirming the harmonization procedure functioned as intended. Given that site accounted for minimal variance even prior to harmonization, all primary analyses reported in the main text use the original (unharmonized) CAP metrics; harmonization is reported here as a robustness check on residual scanner effects rather than as a step in the primary analytic pipeline.

| **CAP** | **RAW SITE R²** | **HARMONIZED SITE R²** |
| --- | --- | --- |
| **DWELL TIME** |  |  |
| AT-SM | 0.0071 | 0.0010 |
| DM | 0.0025 | 0.0005 |
| FP-DM | 0.0054 | 0.0002 |
| VS-SC | 0.0407 | 0.0009 |
| **ENTRIES** |  |  |
| AT-SM | 0.0181 | 0.0004 |
| DM | 0.0227 | 0.0005 |
| FP-DM | 0.0020 | 0.0009 |
| VS-SC | 0.0000 | 0.0001 |

**Site-associated variance in CAP dwell time (DUR) and entries (ENT), before and after longitudinal ComBat harmonization.** R² from linear regression of each metric on site (3-level factor: Amsterdam, Leiden, Groningen), computed on all available pident-wave observations (*N* = 543 observations, all waves and both groups pooled), before and after longitudinal ComBat harmonization (group, age, and sex protected as covariates; random intercept per subject). Harmonized R² ≤ 0.001 across all eight CAP-metric combinations confirms effective removal of residual site variance; the modest magnitude of raw R² indicates minimal site-driven bias in the original (unharmonized) metrics used for all primary analyses.

#### **S5. CAP Extraction and Validation.**

**S5.1 Clustering.** All retained fMRI volumes were entered into a seed-free, data-driven k-means clustering using correlation distance (1 − Pearson r), which groups frames by the shape of their spatial co-activation pattern across parcels while being insensitive to overall signal amplitude (Figure 1B). No activation threshold was applied; both positive (co-activation) and negative (co-deactivation) parcel values were retained. Frames were not preselected on any seed region. Volumes assigned to each cluster were averaged to yield k co-activation pattern (CAP) maps, which were normalized by the within-cluster standard error to produce Z-statistic maps. For quality assessment, we computed a post-hoc dispersion metric (mean correlation between each CAP centroid and its assigned frames) and per-frame correlations to the assigned CAP. CAP temporal metrics were characterized by two measures: dwell time (mean duration of consecutive frames assigned to a given state, in seconds) and entries (number of times a state was initiated; Figure 1A).

**S5.2 Optimal K Selection and Internal Recovery.** Optimal K was selected by evaluating solutions across K = 3–20 using three complementary criteria. **Clustering stability** (Figure 1C): assessed via split-half resampling (consensus index (9)). K = 4 and K = 10 exhibited the highest stability coefficients and were selected as the primary and secondary solutions respectively. **Inter-solution consistency** (Figure 1D; Figure S2A): to verify that the K = 4 spatial patterns are reproducible and not artefacts of a single coarse solution, we assessed their recovery within the finer-grained K = 10 solution. Each Z-statistic map was thresholded (z > 0.1) to produce a binary spatial mask, and pairwise spatial overlap between corresponding K = 4 and K = 10 CAP masks was quantified using the Dice Similarity Coefficient (DSC = 2|A∩B| / (|A| + |B|); the harmonic mean of recall and precision, ranging from 0 to 1), recall (|A∩B| / |A|; the proportion of the K = 4 CAP's active parcels also active in the matched K = 10 CAP, i.e. sensitivity to the reference map), and precision (|A∩B| / |B|; the proportion of the K = 10 CAP's active parcels overlapping the K = 4 CAP, i.e. specificity of the match). Globally, overlap between the K = 4 and K = 10 solutions was 81% across spatial maps (Figure 1D). At the level of individual CAPs, all four K = 4 states were recovered within the K = 10 clustering (see also Figure S2A):

| **CAP** | **K=10 match** | **DSC** | **Recall** | **Precision** | **Union coverage** |
| --- | --- | --- | --- | --- | --- |
| AT-SM | CAP1 | 0.77 | 63.8% | 96.7% | 100% |
| DM | CAP3 | 0.77 | 65.7% | 92.2% | 100% |
| FP-DM | CAP2 | 0.85 | 86.3% | 83.8% | 100% |
| VS-SC | CAP7 | 0.82 | 81.9% | 83.1% | 100% |

The uniformly high precision values confirm the K = 10 matches are spatially focused rather than diffuse, and the 100% union coverage confirms no K = 4 state is split across non-overlapping K = 10 clusters. FP-DM_CAP_ showed the highest internal recovery of the four states (DSC = 0.85). Two representative K = 10 clusters (CAP9, CAP10) that did not correspond to any K = 4 state illustrate the noisy, poorly differentiated patterns that motivated selecting K = 4 over K = 10 as the primary solution (Figure S2A).

**S5.3 Cross-Dataset Replication (NESDA vs. MOTAR).** To evaluate cross-dataset replicability, NESDA CAPs (rows 1–4) were spatially compared against an independent external CAP solution from the MOTAR dataset ((10) K = 5; columns 1–5) using the Dice Similarity Coefficient (Figure 1E; Figure S2B). Because each CAP map contains both positively co-active and negatively co-deactive regions, the spatial comparison was performed separately across four mask combinations — NESDA coactive vs. MOTAR coactive, NESDA coactive vs. MOTAR de-coactive, NESDA de-coactive vs. MOTAR coactive, and NESDA de-coactive vs. MOTAR de-coactive (Figure S2B) — to verify sign-specific spatial correspondence and rule out spurious cross-sign matches. Coactive (positive) and de-coactive (negative) spatial maps showed overall overlap of 89% and 93% respectively (Figure 1E), confirming cross-dataset replicability of the K = 4 solution. At the level of individual CAPs (Figure S2B):

| **CAP** | **Match in MOTAR** | **DSC** |
| --- | --- | --- |
| AT-SM | CAP1 | 0.71 |
| DM | CAP2 | 0.75 |
| FP-DM | CAP3/CAP4* | 0.41 / 0.48 |
| VS-SC | CAP5 | 0.76 |

*NESDA FP-DM_CAP_ matched two CAPs found in the MOTAR dataset (CAP3/CAP4) encompassing two fronto-parietal states. DSC**:** Dice Similarity Coefficient

FP-DM_CAP_ showed moderate overlap with two MOTAR fronto-parietal states rather than one, consistent with the K = 4 solution merging two fronto-parietal configurations that are resolved as separate states at K = 5; correspondingly, FP-DM_CAP_ was the state with the lowest external replication DSC despite the highest internal (K = 10) recovery. VS-SC_CAP_ was the most spatially consistent state across datasets (DSC = 0.76). Off-diagonal and cross-sign cells were uniformly near zero (Figure S2B), confirming sign-specific spatial correspondence and ruling out cross-sign artefacts across independent datasets.

##### **S6. Within-person CAP–symptom coupling: linear mixed models.**

**S6.1 Rationale.** The within-between mixed model separates two quantities a pooled regression would conflate: whether people with higher average symptoms differ in CAP expression (between-person, trait), and whether a person's session-to-session symptom fluctuations track their own CAP fluctuations (within-person). The model also uses all observations, including single-session participants, which inform the between-person and residual estimates (11,12).

**S6.2 Decomposition.** To separate stable individual differences from moment-to-moment change, each symptom score *S* of subject *i* at session *t* was split into two parts. The between-person component (*s_b*) is the subject's own mean across their available sessions, *S_b(i)* = mean of *S* over *i*'s sessions; it captures how symptomatic that person generally is " (i.e., their trait level). . The within-person component (*s_w*) is each session's deviation from that personal mean, *S_w(it)* = *S_it* − *S_b(i)*; it captures whether the person was doing better or worse than usual for them at that one session. Entering both in the same model lets *s_b* absorb stable between-person differences, including unmeasured, time-invariant confounders such as baseline severity or personality. So that the within-person coefficient *s_w* estimates the association free of them. Both components were z-scored (*s_b*, *s_w*). Briefly: a significant s_w means that when a person's symptoms rise above their own average, their CAP expression rises too. s_b, reported alongside, is just how symptomatic that person is overall.

**S6.3 Outcome.** The rank-transformed CAP metric, rank(CAP), ranked across all observations entering each fit, a distribution-robust choice for the bounded, non-normal CAP metrics.

**S6.4 Models.** Two complementary rank-LMMs were fitted per metric (DUR, ENT) and symptom (IDS, BAI, FQ), both including covariates for age, sex, education level, drug use, and scanning site.

**Group-interaction model (LMM: symptom x group) .** Fitted once per CAP × symptom to the combined AFF + CTR sample, testing whether within-person coupling differs between groups:

*rank(CAP)_it = β0 + β1·s_b + β2·s_w + β3·group + β4·(s_b × group) + β5·(s_w × group) + β6·age + β7·sex + β8·site + β9·aedulvl + β10·drugs + u_i + ε_it,*

with *u_i ~ N(0, τ²)*. Group was CTR-referenced, so *β2* = coupling in CTR, *β5* = the *s_w × group interaction* (the AFF-vs-CTR difference, the test of interest), and *β2 + β5* = coupling in AFF. *s_b* and *s_w* were z-scored over the combined AFF + CTR sample so that both groups share a common scaling, which the interaction term requires to be interpretable. Degrees of freedom were Satterthwaite; effect size was partial η²p = t²/(t² + df). Models used REML and permitted singular fits.

**Session-moderated model (LMM: symptom x session) .** Fitted separately within each group (AFF, CTR), testing whether within-person coupling varies across the follow-up:

*rank(CAP)_it = β0 + β1·s_b + β2·s_w + β3·session + β4·(s_w × session) + β5·age + β6·sex + β7·site + β8·aedulvl + β9·drugs + u_i + ε_it,*

with session as a three-level factor (Y0, Y2, Y9; Y0 reference) and *s_b*, *s_w* z-scored within each group's own sample. The primary test is the omnibus *s_w × session* interaction (Kenward-Roger F-test), asking whether coupling strengthens or weakens across the nine-year follow-up. Post-hoc, the session-specific within-person slope (*s_w* at each session) was contrasted pairwise across the three non-overlapping intervals (Y0→Y2, Y0→Y9, Y2→Y9) via estimated marginal trends, each reported with SE, df, and 95% CI. Only subjects with ≥ 2 available sessions were retained, as single-session participants carry no within-person change information for this model.

**S6.5 Diagnostics.** A separate diagnostic analysis tested the assumptions underlying the pooled group-interaction model above — i.e., whether AFF and CTR can be validly combined into a single model rather than requiring group-specific structure. For each CAP × symptom × metric cell, a reduced model (covariates and the s_b/s_w main effects only, no group term) was compared by likelihood-ratio test against the full group-interaction model, testing whether adding group improves fit at all. The shared-residual-variance assumption of the pooled model was assessed with Levene's test on residuals by group, and with a group-specific residual-variance model (nlme, varIdent), compared to the standard model by AIC and likelihood-ratio test. Singular-fit flags and per-group Shapiro–Wilk tests on residuals were additionally reported for each cell (Table S–). These diagnostics do not themselves constitute a reported inferential test; they establish whether the pooled group-interaction model in S5.4 is an appropriate specification.

**S6.6 Correcting multiple testing.** Benjamini–Hochberg FDR was applied separately for CPS metrics (DUR and ENT (independent families)) and separately for each of the two models in S5.4, since they are corrected as distinct families:

- Group-interaction model: FDR across the 4 CAPs × 3 symptoms (12 tests) entering the combined-sample *s_w × group* interaction test, one family per metric (not split by group, since the model itself is fitted on the combined sample).
- Session-moderated model: FDR across the omnibus *s_w × session* interaction p-value for the 4 CAPs × 3 symptoms (12 tests), applied separately within AFF and within CTR (24 tests total per metric, corrected as two independent 12-test families). The post-hoc pairwise interval contrasts (Y0→Y2, Y0→Y9, Y2→Y9) are descriptive follow-up to a significant omnibus interaction and are not themselves included in this FDR family.

All models were implemented in R, lme4/lmerTest (REML; Satterthwaite degrees of freedom for coefficient-level tests, Kenward-Roger for the session-moderated omnibus interaction test); post-hoc interval contrasts used emmeans; diagnostic variance modelling used nlme (varIdent) and car (Levene's test).

#### **S7. Supplementary References.**

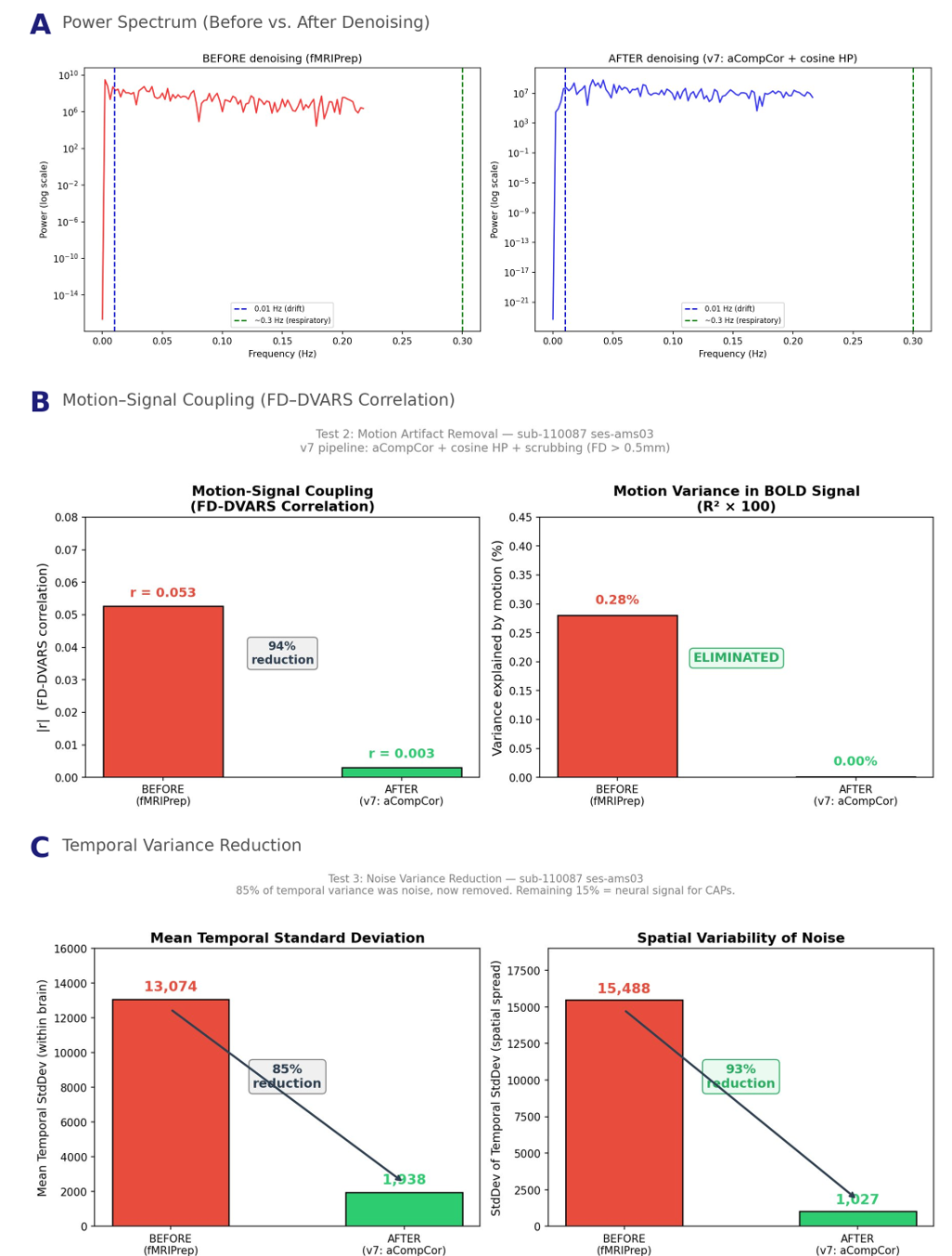

**Supplementary Figure S1. Denoising pipeline validation for a random participant (sub-110087, ses-ams03).** **A.** Power spectrum of the mean brain BOLD timeseries before (red) and after (blue) denoising. The steep drop below 0.01 Hz confirms effective drift removal by cosine regression; the flat plateau from 0.01–0.20 Hz confirms preservation of neural signal. **B.** Motion–signal coupling quantified as the FD–DVARS correlation (left) and motion variance in the BOLD signal (right). After denoising, the correlation drops from r = 0.053 to r = 0.003 (94% reduction), with motion-explained variance eliminated to 0.00%. **C.** Temporal standard deviation (tSD) before and after denoising. Mean tSD was reduced from 13,074 to 1,938 (85% reduction) and spatial variability from 15,488 to 1,027 (93% reduction), both within the expected 70–95% pass range. All three tests confirm that

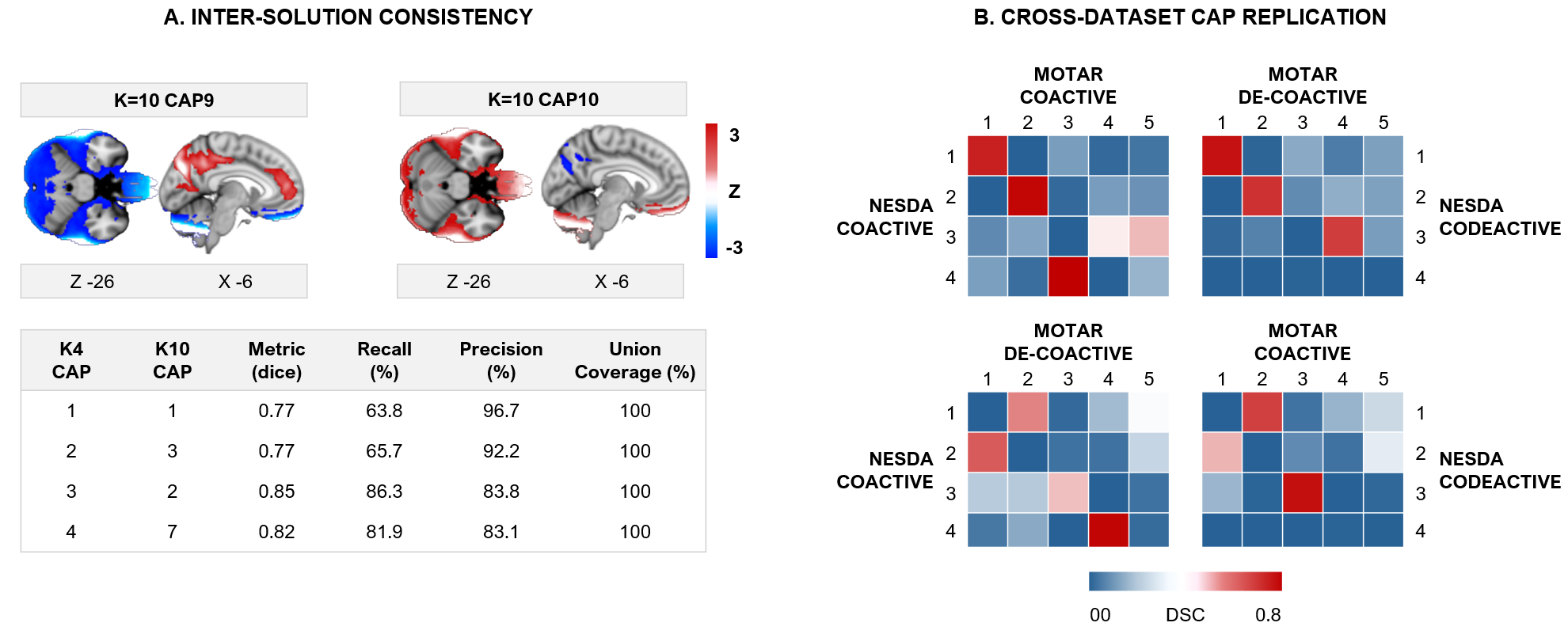

**Figure S2. Inter-clustering solution stability and cross-dataset replication of the NESDA K=4 CAP solution. A.** **Intercluster consistency.** Representative K=10 CAPs illustrating examples of noise-like, poorly differentiated clusters (CAP9 and CAP10; axial slice Z=−26, sagittal slice X=−6; color scale: z-score ±3) that emerge when the number of clusters is over-specified, supporting K=4 as the optimal solution. Nevertheless, despite the presence of such noisy clusters in the K=10 solution, all four K=4 spatial patterns were recovered within the K=10 parcellation, as quantified by Dice coefficient (DSC=0.77–0.85), recall (63.8–86.3%), precision (83.1–96.7%), and union coverage (100% for all four CAPs). **B. External cross-validation.** Dice coefficient matrices quantifying spatial overlap between NESDA K=4 CAPs and MOTAR K=5 CAPs across four mask combinations: coactive–coactive (top left), coactive–de-coactive (top right), de-coactive–coactive (bottom left), and de-coactive–coactive (bottom right). Rows = NESDA CAPs 1–4; columns = MOTAR CAPs 1–5. Color scale: DSC 0.0–0.8. Strong coactive–coactive overlap was observed for NESDA CAP1–MOTAR CAP1 (DSC=0.71), CAP2–MOTAR CAP2 (DSC=0.75), and CAP4–MOTAR CAP5 (DSC=0.76), with NESDA CAP3 showing moderate overlap with two MOTAR fronto-parietal states (MOTAR CAP3/CAP4; DSC=0.41–0.48). Off-diagonal and cross-sign cells were uniformly low, confirming sign-specific spatial correspondence across independent datasets.

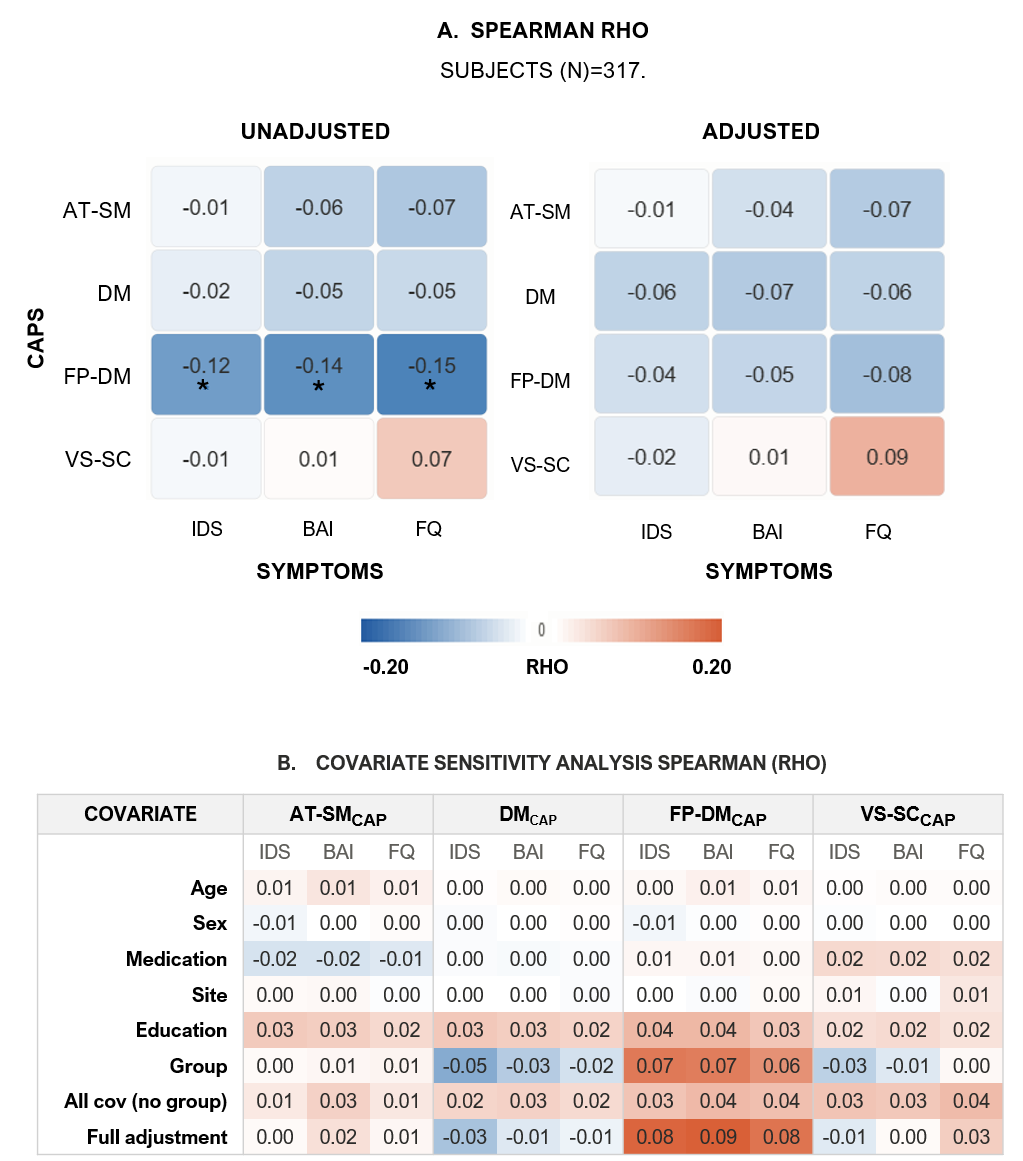

**Supplementary Figure S3.** Covariate sensitivity of baseline CAP–symptom associations and group-driven confounding in the pooled-sample analysis. Spearman correlations (rho) between CAP dwell time and symptom severity (IDS, BAI, FQ) in the full baseline sample (N = 317), shown unadjusted (left) and fully adjusted (right; covariates: age, sex, medication, site, education, group). Bottom heatmaps show the change in each estimate (Δ) relative to the unadjusted model as covariates are added sequentially; effects were minimal except for group membership (AFF vs. CTR), which produced the largest shifts across CAP–symptom cells. Pooling AFF and CTR participants obscured within-group associations: the AT-SM_CAP–IDS effect was significant in CTR but absent in the pooled sample, and FP-DM_CAP–symptom associations were entirely attributable to group composition — consistent with Simpson's paradox (13). These results support the group-stratified approach in Figure 3. Abbreviations: AFF, affective group; BAI, Beck Anxiety Inventory; CAP, co-activation pattern; CTR, control group; FQ, Fear Questionnaire; IDS, Inventory of Depressive Symptomatology. **q* < 0.05.

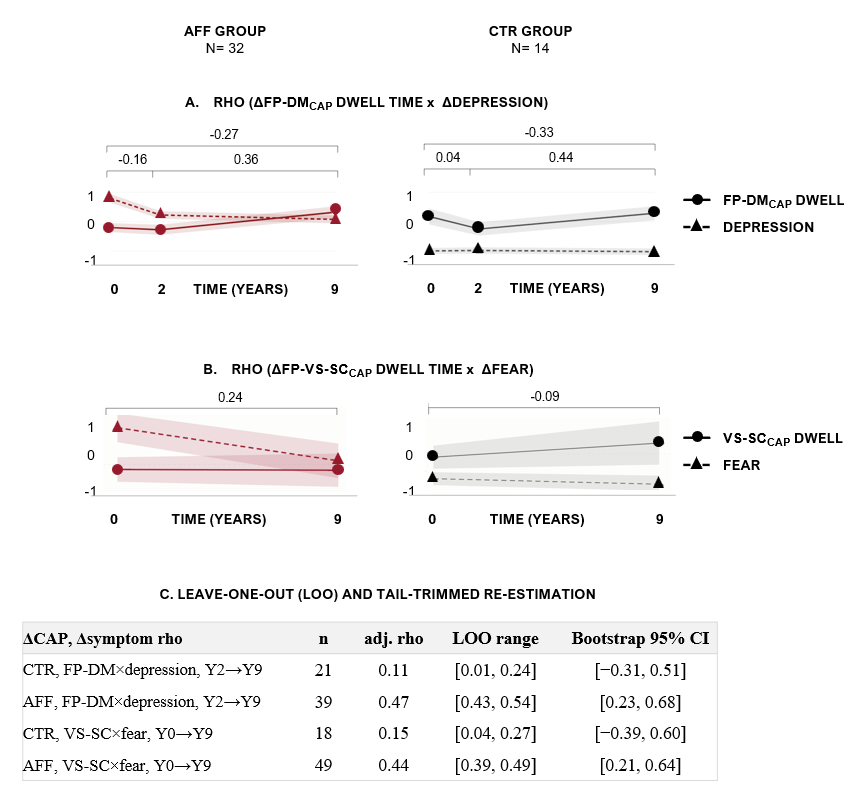

**Supplementary Figure S4. Sensitivity analysis: replication of main findings in participants with complete three-wave data.** Change–change correlations (Spearman rho), restricted to participants with fMRI data at all three waves (Y0, Y2, Y9; total N = 46: AFF n = 32, CTR n = 14). Trajectories show z-scored group means (±1 SE); brackets and rho values indicate fully adjusted Spearman correlations for each interval. Left column: AFF (dark red); right column: CTR (grey). **A.** FP-DM_CAP_ dwell time × depression severity. The positive AFF coupling over Y2→Y9 (rho = +0.36) replicates the direction and approximate magnitude of the main finding (rho = +0.47), consistent with robustness to sample composition; no coupling is observed in CTR. **B.** VS-SC_CAP_ dwell time × fear severity (Y0→Y9). The positive direction in AFF is maintained (rho = +0.24) but does not reach significance, reflecting the expected loss of power in the reduced sample (n = 32 vs. n = 49 in the main analysis); no coupling is observed in CTR. **C.** Leave-one-out (LOO) and tail-trimmed re-estimation. In **AFF**, both couplings are robust to individual participants and tail composition: LOO ranges stay narrow and clear of zero (FP-DM_CAP_ × depression: [0.43, 0.54]; VS-SC_CAP_ × fear: [0.39, 0.49]), rho remains positive under all trims (5–15%), and bootstrap 95% CIs exclude zero for both ([0.23, 0.68]; [0.21, 0.64]). In **CTR**, VS-SC_CAP_ × fear is a stable null (LOO [0.04, 0.27], positive under all trims), whereas FP-DM_CAP_ × depression, though LOO-stable ([0.01, 0.24]), turns negative under 10–15% trimming — an underpowered near-zero effect rather than a robust null. Wide bootstrap CIs in CTR ([−0.31, 0.51]; [−0.39, 0.60]) reflect limited precision (n = 18–21), not instability. Together, panels A–C show the AFF findings are not driven by influential participants, support the directionality and outlier-robustness of the FP-DM_CAP_ × depression coupling, and confirm the VS-SC_CAP_ × fear association is power-limited rather than spurious.

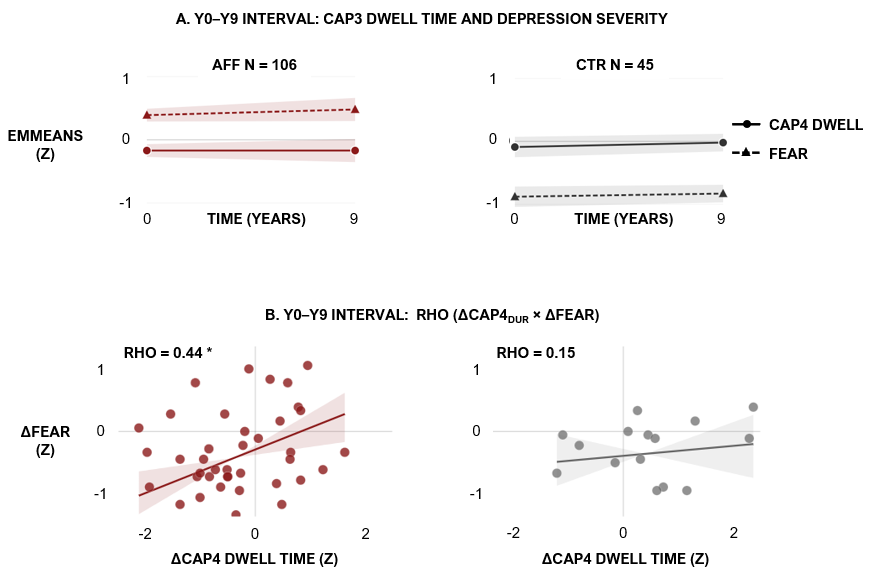

**Supplementary Figure S5. Group-mean trajectories and within-person change–change coupling for VS-SC_CAP_ dwell time and fear severity over the Y0→Y9 interval. (A)** Group-mean trajectories. Estimated marginal means (±1 SE; z-scored) of VS-SC_CAP_ dwell time (solid line, circles) and fear severity (FQ; dashed line, triangles) at baseline (Y0) and 9-year follow-up (Y9), in affective patients (AFF, dark red; n = 106) and healthy controls (CTR, grey; n = 45), derived from LMM. VS-SC_CAP_ dwell time was essentially flat across 9 years in both groups (AFF: −0.17→−0.17; CTR: −0.09→−0.02). Fear severity remained persistently elevated in AFF (EMM: +0.39→+0.48) and consistently low in CTR (−0.88→−0.83). The absence of co-variation between the two trajectories indicates no meaningful group-level coupling over this interval. **B.** Within-person change–change coupling. Each point represents one subject. X-axis: within-person change in VS-SC_CAP_ dwell time (Δz) from Y0 to Y9; y-axis: within-person change in fear severity (Δz) over the same interval. AFF: dark red; CTR: grey. Regression line derived from the fully adjusted Spearman rho (covariates: age, sex, drug use, recruitment site, education level), anchored at the data centroid. Shaded band: 95% confidence interval (Fisher Z-transformation). Axes fixed at ±1. Reported rho reflects the complete interval sample (AFF n = 49, CTR n = 18). Despite the flat group-mean trajectories in Panel A, Panel B reveals a significant positive within-person coupling in AFF (rho = +0.44, q < 0.05): individuals who maintained or increased VS-SC_CAP_ dwell time over 9 years retained more fear symptoms, while those whose dwell time declined showed less fear scoring (i.e., fear improvement). This coupling was absent in CTR (rho = +0.15, q > 0.38). The contrast between panels illustrates that the association is entirely a within-person phenomenon — invisible at the group-mean level yet robust across individuals in the affective group. Values from unadjusted modelling are reported in table S15.* *q* < 0.05; FDR-corrected across all CAP × symptom × interval combinations within each group (36 tests).

| **AT-SM_CAP_ CO-ACTIVATION** | | | | | |
| --- | --- | --- | --- | --- | --- |
| **Region_AAL3** | **Hemi** | **Voxels** | **% Total** | **Yeo17_Network** | **Functional System** |
| **Cluster 1 \| 22,144 voxels \| peak z = +3.20 \| Mid Cingulate / Sensorimotor** | | | | | |
| Postcentral_R | R | 1528 | 6.6 | SomMotA | Sensorimotor |
| SupraMarginal_R | R | 1332 | 5.7 | SomMotB | Sensorimotor |
| Insula_R | R | 1236 | 5.3 | SalVentAttnA | Salience |
| Temporal_Sup_L | L | 1145 | 4.9 | TempPar | Temporoparietal |
| Insula_L | L | 1094 | 4.7 | SalVentAttnA | Salience |
| Supp_Motor_Area_R | R | 1037 | 4.5 | SomMotB | Sensorimotor |
| Rolandic_Oper_R | R | 1004 | 4.3 | SomMotB | Sensorimotor |
| Precentral_R | R | 992 | 4.3 | SomMotA | Sensorimotor |
| Postcentral_L | L | 988 | 4.3 | SomMotA | Sensorimotor |
| Cingulate_Mid_R | R | 881 | 3.8 | SalVentAttnB | Salience |
| SupraMarginal_L | L | 880 | 3.8 | SomMotB | Sensorimotor |
| Supp_Motor_Area_L | L | 813 | 3.5 | SomMotB | Sensorimotor |
| Rolandic_Oper_L | L | 768 | 3.3 | SomMotB | Sensorimotor |
| Temporal_Sup_R | R | 744 | 3.2 | TempPar | Temporoparietal |
| Cingulate_Mid_L | L | 728 | 3.1 | SalVentAttnB | Salience |
| Frontal_Inf_Oper_R | R | 627 | 2.7 | ContC | Control |
| Frontal_Sup_2_R | R | 590 | 2.5 | ContA | Control |
| Parietal_Inf_L | L | 576 | 2.5 | ContB | Control |
| Precentral_L | L | 454 | 2.0 | SomMotA | Sensorimotor |
| Frontal_Inf_Oper_L | L | 398 | 1.7 | ContC | Control |
| Parietal_Sup_L | L | 394 | 1.7 | DorsAttnB | Attention |
| Parietal_Sup_R | R | 339 | 1.5 | DorsAttnB | Attention |
| Parietal_Inf_R | R | 323 | 1.4 | ContB | Control |
| Frontal_Sup_2_L | L | 270 | 1.2 | ContA | Control |
| Precuneus_R | R | 234 | 1.0 | DefaultC | DefaultMode |
| Precuneus_L | L | 195 | 0.8 | DefaultC | DefaultMode |
| Frontal_Mid_2_R | R | 168 | 0.7 | ContA | Control |
| Frontal_Inf_Tri_R | R | 122 | 0.5 | ContA | Control |
| ACC_sup_L | L | 71 | 0.3 | SalVentAttnA | Salience |
| Frontal_Inf_Tri_L | L | 68 | 0.3 | ContA | Control |
| **Cluster 4 \| 250 voxels \| peak z = +1.65 \| IFG / MFG Right** | | | | | |
| Frontal_Mid_2_R | R | 130 | 0.6 | ContA | Control |
| Frontal_Inf_Tri_R | R | 120 | 0.5 | ContA | Control |
| **Cluster 5 \| 408 voxels \| peak z = +1.77 \| MFG Left** | | | | | |
| Frontal_Mid_2_L | L | 404 | 1.7 | ContA | Control |
| **Cluster 6 \| 426 voxels \| peak z = +1.89 \| MFG Right** | | | | | |
| Frontal_Mid_2_R | R | 372 | 1.6 | ContA | Control |

| **AT-SM_CAP_ CO-DEACTIVATION** | | | | | |
| --- | --- | --- | --- | --- | --- |
| **Region_AAL3** | **Hemi** | **Voxels** | **% Total** | **Yeo17_Network** | **Functional System** |
| **Cluster 5 \| 9,739 voxels \| peak z = −2.86 \| Medial Prefrontal (mPFC)** | | | | | |
| Frontal_Sup_2_L | L | 1648 | 7.5 | ContA | Control |
| Frontal_Sup_Medial_L | L | 1540 | 7.0 | DefaultB | DefaultMode |
| Frontal_Sup_2_R | R | 1337 | 6.0 | ContA | Control |
| Frontal_Sup_Medial_R | R | 1293 | 5.8 | DefaultB | DefaultMode |
| Frontal_Mid_2_L | L | 1219 | 5.5 | ContA | Control |
| Frontal_Mid_2_R | R | 582 | 2.6 | ContA | Control |
| Frontal_Med_Orb_L | L | 467 | 2.1 | DefaultB | DefaultMode |
| Frontal_Med_Orb_R | R | 422 | 1.9 | DefaultB | DefaultMode |
| ACC_pre_L | L | 262 | 1.2 | DefaultB | DefaultMode |
| ACC_pre_R | R | 208 | 0.9 | DefaultB | DefaultMode |
| **Cluster 6 \| 5,111 voxels \| peak z = −3.53 \| Precuneus / PCC** | | | | | |
| Precuneus_L | L | 1307 | 5.9 | DefaultC | DefaultMode |
| Precuneus_R | R | 1162 | 5.3 | DefaultC | DefaultMode |
| Calcarine_L | L | 400 | 1.8 | VisCent | Visual |
| Cuneus_L | L | 347 | 1.6 | VisPeri | Visual |
| Cingulate_Mid_L | L | 319 | 1.4 | SalVentAttnB | Salience |
| Cingulate_Mid_R | R | 301 | 1.4 | SalVentAttnB | Salience |
| Cingulate_Post_L | L | 277 | 1.3 | DefaultA | DefaultMode |
| Calcarine_R | R | 193 | 0.9 | VisCent | Visual |
| Cuneus_R | R | 142 | 0.6 | VisPeri | Visual |
| Cingulate_Post_R | R | 135 | 0.6 | DefaultA | DefaultMode |
| Lingual_R | R | 123 | 0.6 | VisCent | Visual |
| **Cluster 7 \| 1,965 voxels \| peak z = −3.21 \| Angular Gyrus Right** | | | | | |
| Angular_R | R | 1235 | 5.6 | DefaultA | DefaultMode |
| Parietal_Inf_R | R | 272 | 1.2 | ContB | Control |
| Temporal_Mid_R | R | 170 | 0.8 | DefaultA | DefaultMode |
| Occipital_Mid_R | R | 113 | 0.5 | VisPeri | Visual |
| Parietal_Sup_R | R | 71 | 0.3 | DorsAttnB | Attention |
| **Cluster 8 \| 2,729 voxels \| peak z = −3.38 \| Angular Gyrus Left** | | | | | |
| Angular_L | L | 1046 | 4.7 | DefaultA | DefaultMode |
| Parietal_Inf_L | L | 499 | 2.3 | ContB | Control |
| Occipital_Mid_L | L | 330 | 1.5 | VisPeri | Visual |
| Temporal_Mid_L | L | 208 | 0.9 | DefaultA | DefaultMode |
| Parietal_Sup_L | L | 151 | 0.7 | DorsAttnB | Attention |
| **Cluster 3 \| 1,006 voxels \| peak z = −1.96 \| Middle Temporal Gyrus Right** | | | | | |
| Temporal_Mid_R | R | 896 | 4.1 | DefaultA | DefaultMode |
| **Cluster 4 \| 1,558 voxels \| peak z = −2.13 \| Middle Temporal Gyrus Left** | | | | | |
| Temporal_Mid_L | L | 1475 | 6.7 | DefaultA | DefaultMode |

**Supplementary Table S1. AT-SM_CAP_ (Attention-Sensorimotor) co-activation and de-activation pattern: Regional composition and functional network assignment.** Anatomical regions comprising the AT-SM_CAP_ co-activation pattern (z ≥ +1.0, cluster ≥ 50 voxels; co-activation: 22,676 total voxels; 4 clusters; de-activation: 22,108 total voxels; 8 clusters, of which 6 ≥ 50 voxels shown) and the AT-SM_CAP_ de-activation pattern (z ≤ −1.0, cluster ≥ 50 voxels), predominantly comprising Default Mode Network regions. Regions with ≥ 61 voxels are reported. Region_AAL3: Automated Anatomical Labelling Atlas v3.1 nomenclature. Hemi: L, left; R, right. Voxels: 2 mm³ MNI space. % Total: proportion of all voxels in the respective pattern. Yeo17_Network: assignment to Yeo's 17-network parcellation (Schaefer et al., 2018). Functional System: higher-level cognitive system label.

| **DM_CAP_ COACTIVATION** | | | | | | | | | | | | | | | | | | | |
| --- | --- | --- | --- | --- | --- | --- | --- | --- | --- | --- | --- | --- | --- | --- | --- | --- | --- | --- | --- |
| **Region_AAL3** | | **Hemi** | | | **Voxels** | | | | | **% Total** | | | | **Yeo17_Network** | | | | **Functional System** | |
| **Cluster 7 \| 9,892 voxels \| peak z = +3.34 \| Medial Prefrontal (mPFC) / vmPFC** | | | | | | | | | | | | | | | | | | | |
| Frontal_Sup_Medial_L | | L | | | 1675 | | | | | 6.2 | | | | DefaultB | | | | DefaultMode | |
| Frontal_Sup_2_L | | L | | | 1522 | | | | | 5.6 | | | | DefaultB | | | | DefaultMode | |
| Frontal_Sup_Medial_R | | R | | | 1378 | | | | | 5.1 | | | | DefaultB | | | | DefaultMode | |
| Frontal_Sup_2_R | | R | | | 979 | | | | | 3.6 | | | | DefaultB | | | | DefaultMode | |
| Frontal_Med_Orb_R | | R | | | 634 | | | | | 2.3 | | | | DefaultB | | | | DefaultMode | |
| Frontal_Med_Orb_L | | L | | | 558 | | | | | 2.0 | | | | DefaultB | | | | DefaultMode | |
| Frontal_Mid_2_L | | L | | | 549 | | | | | 2.0 | | | | ContA | | | | Control | |
| ACC_pre_L | | L | | | 485 | | | | | 1.8 | | | | DefaultB | | | | DefaultMode | |
| ACC_pre_R | | R | | | 364 | | | | | 1.3 | | | | DefaultB | | | | DefaultMode | |
| Rectus_L | | L | | | 251 | | | | | 0.9 | | | | DefaultB | | | | DefaultMode | |
| Rectus_R | | R | | | 194 | | | | | 0.7 | | | | DefaultB | | | | DefaultMode | |
| Olfactory_L | | L | | | 152 | | | | | 0.6 | | | | DefaultB | | | | DefaultMode | |
| Olfactory_R | | R | | | 94 | | | | | 0.3 | | | | DefaultB | | | | DefaultMode | |
| ACC_sub_L | | L | | | 92 | | | | | 0.3 | | | | DefaultB | | | | DefaultMode | |
| **Cluster 9 \| 3,319 voxels \| peak z = +3.39 \| Precuneus / PCC** | | | | | | | | | | | | | | | | | | | |
| Precuneus_L | | L | | | 890 | | | | | 3.3 | | | | DefaultC | | | | DefaultMode | |
| Precuneus_R | | R | | | 689 | | | | | 2.5 | | | | DefaultC | | | | DefaultMode | |
| Cingulate_Post_L | | L | | | 241 | | | | | 0.9 | | | | DefaultA | | | | DefaultMode | |
| Cingulate_Mid_L | | L | | | 222 | | | | | 0.8 | | | | SalVentAttnB | | | | Salience | |
| Cingulate_Mid_R | | R | | | 203 | | | | | 0.7 | | | | SalVentAttnB | | | | Salience | |
| Calcarine_L | | L | | | 330 | | | | | 1.2 | | | | VisCent | | | | Visual | |
| Cuneus_L | | L | | | 188 | | | | | 0.7 | | | | VisPeri | | | | Visual | |
| Cingulate_Post_R | | R | | | 104 | | | | | 0.4 | | | | DefaultA | | | | DefaultMode | |
| **Cluster 4 \| 5,199 voxels \| peak z = +2.86 \| Temporal Pole / MTG / OFC Left** | | | | | | | | | | | | | | | | | | | |
| Temporal_Mid_L | | L | | | 2023 | | | | | 7.4 | | | | DefaultA | | | | DefaultMode | |
| Temporal_Inf_L | | L | | | 732 | | | | | 2.7 | | | | DefaultA | | | | DefaultMode | |
| Temporal_Pole_Mid_L | | L | | | 487 | | | | | 1.8 | | | | LimbicA | | | | Limbic | |
| Temporal_Pole_Sup_L | | L | | | 383 | | | | | 1.4 | | | | LimbicA | | | | Limbic | |
| Frontal_Inf_Orb_2_L | | L | | | 289 | | | | | 1.1 | | | | ContB | | | | Control | |
| OFCpost_L | | L | | | 229 | | | | | 0.8 | | | | LimbicB | | | | Limbic | |
| Frontal_Inf_Tri_L | | L | | | 140 | | | | | 0.5 | | | | ContA | | | | Control | |
| OFClat_L | | L | | | 136 | | | | | 0.5 | | | | LimbicB | | | | Limbic | |
| Insula_L | | L | | | 75 | | | | | 0.3 | | | | SalVentAttnA | | | | Salience | |
| **Cluster 2 \| 3,633 voxels \| peak z = +2.48 \| Temporal Pole / MTG / OFC Right** | | | | | | | | | | | | | | | | | | | |
| Temporal_Mid_R | | R | | | 1083 | | | | | 4.0 | | | | DefaultA | | | | DefaultMode | |
| Temporal_Inf_R | | R | | | 724 | | | | | 2.7 | | | | DefaultA | | | | DefaultMode | |
| Temporal_Pole_Mid_R | | R | | | 664 | | | | | 2.4 | | | | LimbicA | | | | Limbic | |
| Temporal_Pole_Sup_R | | R | | | 263 | | | | | 1.0 | | | | LimbicA | | | | Limbic | |
| OFCpost_R | | R | | | 233 | | | | | 0.9 | | | | LimbicB | | | | Limbic | |
| OFClat_R | | R | | | 94 | | | | | 0.3 | | | | LimbicB | | | | Limbic | |
| Frontal_Inf_Orb_2_R | | R | | | 86 | | | | | 0.3 | | | | ContB | | | | Control | |
| Insula_R | | R | | | 73 | | | | | 0.3 | | | | SalVentAttnA | | | | Salience | |
| **Cluster 10 \| 1,760 voxels \| peak z = +3.05 \| Angular Gyrus Left** | | | | | | | | | | | | | | | | | | | |
| Angular_L | | L | | | 956 | | | | | 3.5 | | | | DefaultA | | | | DefaultMode | |
| Temporal_Mid_L | | L | | | 208 | | | | | 0.8 | | | | DefaultA | | | | DefaultMode | |
| Occipital_Mid_L | | L | | | 128 | | | | | 0.5 | | | | VisPeri | | | | Visual | |
| Parietal_Inf_L | | L | | | 113 | | | | | 0.4 | | | | DorsAttnB | | | | DorsalAttention | |
| **Cluster 11 \| 1,020 voxels \| peak z = +2.55 \| Angular Gyrus Right** | | | | | | | | | | | | | | | | | | | |
| Angular_R | | R | | | 777 | | | | | 2.9 | | | | DefaultA | | | | DefaultMode | |
| Temporal_Mid_R | | R | | | 117 | | | | | 0.4 | | | | DefaultA | | | | DefaultMode | |
|  | **Cluster 6 \| 283 voxels \| peak z = +1.63 \| Hippocampus / Parahippocampal Left** | | | | | | | | | | | | | | | | | | |
|  | Hippocampus_L | | L | | | 169 | | | | | 0.6 | | | | DefaultA | | | | DefaultMode |
|  | ParaHippocampal_L | | L | | | 82 | | | | | 0.3 | | | | DefaultA | | | | DefaultMode |
|  | **Cluster 8 \| 191 voxels \| peak z = +1.48 \| Hippocampus / Parahippocampal Right** | | | | | | | | | | | | | | | | | | |
|  | Hippocampus_R | | R | | | 89 | | | | | 0.3 | | | | DefaultA | | | | DefaultMode |
|  | ParaHippocampal_R | | R | | | 84 | | | | | 0.3 | | | | DefaultA | | | | DefaultMode |
|  | **Cluster 3 \| 1,008 voxels \| peak z = +1.92 \| Cerebellum Crus1/2 Right** | | | | | | | | | | | | | | | | | | |
|  | Cerebellum_Crus1_R | | R | | | | 536 | 2.0 | | | | Subcortical | | | | Cerebellum | | | |
|  | Cerebellum_Crus2_R | | R | | | | 472 | 1.7 | | | | Subcortical | | | | Cerebellum | | | |
|  | **Cluster 5 \| 503 voxels \| peak z = +1.79 \| Cerebellum Crus2/1 Left** | | | | | | | | | | | | | | | | | | |
|  | Cerebellum_Crus2_L | | L | 321 | | | | | 1.2 | | | | Subcortical | | | | Cerebellum | | |
|  | Cerebellum_Crus1_L | | L | 173 | | | | | 0.6 | | | | Subcortical | | | | Cerebellum | | |
|  | **Cluster 1 \| 422 voxels \| peak z = +1.57 \| Cerebellum 9 / Vermis** | | | | | | | | | | | | | | | | | | |
|  | Cerebellum_9_R | | R | 140 | | | | | 0.5 | | | | Subcortical | | | | Cerebellum | | |
|  | Cerebellum_9_L | | L | 119 | | | | | 0.4 | | | | Subcortical | | | | Cerebellum | | |
|  | Vermis_9 | | B | 77 | | | | | 0.3 | | | | Subcortical | | | | Cerebellum | | |

| **DM_CAP_ CO-DEACTIVATION Dorsal/Ventral Attention + Control** | | | | | | | | | | | | | | | | | | |
| --- | --- | --- | --- | --- | --- | --- | --- | --- | --- | --- | --- | --- | --- | --- | --- | --- | --- | --- |
| **Region_AAL3** | | **Hemi** | | | | **Voxels** | | | | **% Total** | | | | **Yeo17_Network** | | | | **Functional System** |
| **Cluster 4 \| 7,322 voxels \| peak z = −2.72 \| Inferior / Superior Parietal Left** | | | | | | | | | | | | | | | | | | |
| Parietal_Inf_L | L | | | 1509 | | | 6.7 | | | | | DorsAttnB | | | DorsalAttention | | | |
| Parietal_Sup_L | L | | | 1387 | | | 6.2 | | | | | DorsAttnB | | | DorsalAttention | | | |
| SupraMarginal_L | L | | | 731 | | | 3.3 | | | | | SomMotB | | | Sensorimotor | | | |
| Precuneus_L | L | | | 714 | | | 3.2 | | | | | DefaultC | | | DefaultMode | | | |
| Postcentral_L | L | | | 442 | | | 2.0 | | | | | SomMotA | | | Sensorimotor | | | |
| Insula_L | L | | | 422 | | | 1.9 | | | | | SalVentAttnA | | | Salience | | | |
| Frontal_Inf_Oper_L | L | | | 337 | | | 1.5 | | | | | SalVentAttnA | | | Salience | | | |
| Precentral_L | L | | | 336 | | | 1.5 | | | | | SomMotA | | | Sensorimotor | | | |
| Temporal_Sup_L | L | | | 283 | | | 1.3 | | | | | TempPar | | | Temporoparietal | | | |
| Rolandic_Oper_L | L | | | 178 | | | 0.8 | | | | | SomMotB | | | Sensorimotor | | | |
| Occipital_Mid_L | L | | | 124 | | | 0.6 | | | | | VisPeri | | | Visual | | | |
| **Cluster 7 \| 6,807 voxels \| peak z = −2.95 \| Inferior / Superior Parietal Right** | | | | | | | | | | | | | | | | | | |
| Parietal_Sup_R | R | | | 1526 | | | 6.8 | | | | | DorsAttnB | | | DorsalAttention | | | |
| SupraMarginal_R | R | | | 1392 | | | 6.2 | | | | | SomMotB | | | Sensorimotor | | | |
| Postcentral_R | R | | | 780 | | | 3.5 | | | | | SomMotA | | | Sensorimotor | | | |
| Precuneus_R | R | | | 873 | | | 3.9 | | | | | DefaultC | | | DefaultMode | | | |
| Parietal_Inf_R | R | | | 841 | | | 3.7 | | | | | DorsAttnB | | | DorsalAttention | | | |
| Occipital_Mid_R | R | | | 231 | | | 1.0 | | | | | VisPeri | | | Visual | | | |
| Cingulate_Mid_R | R | | | 216 | | | 1.0 | | | | | SalVentAttnB | | | Salience | | | |
| Angular_R | R | | | 175 | | | 0.8 | | | | | DefaultA | | | DefaultMode | | | |
| Occipital_Sup_R | R | | | 189 | | | 0.8 | | | | | DorsAttnB | | | DorsalAttention | | | |
| Temporal_Sup_R | R | | | 129 | | | 0.6 | | | | | TempPar | | | Temporoparietal | | | |
| **Cluster 3 \| 3,002 voxels \| peak z = −2.30 \| IFG / MFG / Insula Right** | | | | | | | | | | | | | | | | | | |
| Frontal_Inf_Oper_R | R | | | 688 | | | 3.1 | | | | | SalVentAttnA | | | Salience | | | |
| Frontal_Sup_2_R | R | | | 591 | | | 2.6 | | | | | DefaultB | | | DefaultMode | | | |
| Insula_R | R | | | 485 | | | 2.2 | | | | | SalVentAttnA | | | Salience | | | |
| Precentral_R | R | | | 470 | | | 2.1 | | | | | SomMotA | | | Sensorimotor | | | |
| Frontal_Mid_2_R | R | | | 258 | | | 1.1 | | | | | ContA | | | Control | | | |
| Rolandic_Oper_R | R | | | 208 | | | 0.9 | | | | | SomMotB | | | Sensorimotor | | | |
| Frontal_Inf_Tri_R | R | | | 66 | | | 0.3 | | | | | ContA | | | Control | | | |
| **Cluster 5 \| 1,472 voxels \| peak z = −2.34 \| MFG / IFG Right (lateral PFC)** | | | | | | | | | | | | | | | | | | |
| Frontal_Mid_2_R | R | | | 1215 | | | 5.4 | | | | | ContA | | | Control | | | |
| Frontal_Inf_Tri_R | R | | | 200 | | | 0.9 | | | | | ContA | | | Control | | | |
| **Cluster 6 \| 918 voxels \| peak z = −2.05 \| MFG / IFG Left (lateral PFC)** | | | | | | | | | | | | | | | | | | |
| Frontal_Mid_2_L | L | | | 698 | | | 3.1 | | | | | ContA | | | Control | | | |
| Frontal_Inf_Tri_L | L | | | 207 | | | 0.9 | | | | | ContA | | | Control | | | |
| **Cluster 8 \| 1,389 voxels \| peak z = −1.98 \| SMA / Mid Cingulate (bilateral)** | | | | | | | | | | | | | | | | | | |
| Supp_Motor_Area_R | R | | | 380 | | | 1.7 | | | | | SomMotB | | | Sensorimotor | | | |
| Cingulate_Mid_R | R | | | 339 | | | 1.5 | | | | | SalVentAttnB | | | Salience | | | |
| Cingulate_Mid_L | L | | | 327 | | | 1.5 | | | | | SalVentAttnB | | | Salience | | | |
| Supp_Motor_Area_L | L | | | 298 | | | 1.3 | | | | | SomMotB | | | Sensorimotor | | | |
| **Cluster 9 \| 546 voxels \| peak z = −1.57 \| MFG / Precentral Left (superior)** | | | | | | | | | | | | | | | | | | |
| Frontal_Sup_2_L | L | | | | 323 | | | 1.4 | | | | | DefaultB | | | | DefaultMode | |
| Precentral_L | L | | | | 153 | | | 0.7 | | | | | SomMotA | | | | Sensorimotor | |
| Frontal_Mid_2_L | L | | | | 63 | | | 0.3 | | | | | ContA | | | | Control | |
| **Cluster 1 \| 468 voxels \| peak z = −1.85 \| Middle / Inferior Temporal Right (posterior)** | | | | | | | | | | | | | | | | | | |
| Temporal_Inf_R | R | | 268 | | | | | | 1.2 | | DefaultA | | | | | DefaultMode | | |
| Temporal_Mid_R | R | | 200 | | | | | | 0.9 | | DefaultA | | | | | DefaultMode | | |
| **Cluster 2 \| 537 voxels \| peak z = −1.89 \| Middle / Inferior Temporal Left (posterior)** | | | | | | | | | | | | | | | | | | |
| Temporal_Mid_L | L | | 242 | | | | | | 1.1 | | DefaultA | | | | | DefaultMode | | |
| Temporal_Inf_L | L | | 145 | | | | | | 0.6 | | DefaultA | | | | | DefaultMode | | |

**Supplementary Table S2. DM_CAP_ co-activation and de-activation pattern: Regional composition and functional network assignment.** Anatomical regions comprising the DM_CAP_ co-activation pattern (z ≥ +1.0, cluster ≥ 50 voxels; **co-activation:** 27,230 total voxels; 11 clusters, dominated by Default Mode Network [60.1%]) and de-activation pattern (z ≤ −1.0, cluster ≥ 50 voxels; de-coactivation: 22,461 total voxels; 9 clusters, dominated by Dorsal Attention [33.0%] and Ventral Attention / Salience [27.3%] networks). Regions with > 60 voxels are reported. Region_AAL3: Automated Anatomical Labelling Atlas v3.1 nomenclature. Hemi: L, left; R, right; B, bilateral. Voxels: 2 mm³ MNI space. % Total: proportion of all voxels in the respective pattern. Yeo17_Network: assignment to Yeo's 17-network parcellation (Schaefer et al., 2018). Functional System: higher-level cognitive system label.

| **FP-DM_CAP_ COACTIVATION** | | | | | | | |
| --- | --- | --- | --- | --- | --- | --- | --- |
| **Region_AAL3** | **Hemi** | **Voxels** | **% Total** | **Yeo17_Network** | | | **Functional System** |
| **Cluster 6 \| 8,262 voxels \| peak z = +2.55 \| MFG / Lateral PFC Right (Control)** | | | | | | | |
| Frontal_Mid_2_R | R | 3141 | 13.8 | ContA | | | Control |
| Frontal_Sup_2_R | R | 1742 | 7.7 | ContA | | | Control |
| Frontal_Sup_Medial_L | L | 616 | 2.7 | DefaultB | | | DefaultMode |
| Frontal_Sup_Medial_R | R | 563 | 2.5 | DefaultB | | | DefaultMode |
| Frontal_Inf_Tri_R | R | 675 | 3.0 | ContA | | | Control |
| Frontal_Inf_Oper_R | R | 430 | 1.9 | ContC | | | Control |
| Precentral_R | R | 130 | 0.6 | SomMotA | | | Sensorimotor |
| Frontal_Inf_Orb_2_R | R | 233 | 1.0 | ContB | | | Control |
| OFCant_R | R | 67 | 0.3 | LimbicB | | | Limbic |
| Cingulate_Mid_R | R | 97 | 0.4 | SalVentAttnB | | | Salience |
| ACC_pre_R | R | 65 | 0.3 | DefaultB | | | DefaultMode |
| **Cluster 7 \| 4,375 voxels \| peak z = +2.13 \| MFG / Lateral PFC Left (Control)** | | | | | | | |
| Frontal_Mid_2_L | L | 2259 | 9.9 | ContA | | | Control |
| Frontal_Inf_Tri_L | L | 847 | 3.7 | ContA | | | Control |
| Frontal_Sup_2_L | L | 520 | 2.3 | ContA | | | Control |
| Precentral_L | L | 325 | 1.4 | SomMotA | | | Sensorimotor |
| Frontal_Inf_Oper_L | L | 143 | 0.6 | ContC | | | Control |
| Frontal_Inf_Orb_2_L | L | 114 | 0.5 | ContB | | | Control |
| **Cluster 9 \| 3,304 voxels \| peak z = +3.08 \| Inferior / Superior Parietal Right** | | | | | | | |
| Angular_R | R | 1235 | 5.4 | DefaultA | | | DefaultMode |
| Parietal_Inf_R | R | 1058 | 4.7 | DorsAttnB | | | DorsalAttention |
| SupraMarginal_R | R | 501 | 2.2 | SomMotB | | | Sensorimotor |
| Parietal_Sup_R | R | 260 | 1.1 | DorsAttnB | | | DorsalAttention |
| Occipital_Mid_R | R | 71 | 0.3 | VisPeri | | | Visual |
| **Cluster 10 \| 3,133 voxels \| peak z = +2.87 \| Inferior / Superior Parietal Left** | | | | | | | |
| Parietal_Inf_L | L | 1521 | 6.7 | DorsAttnB | | | DorsalAttention |
| Angular_L | L | 576 | 2.5 | DefaultA | | | DefaultMode |
| Parietal_Sup_L | L | 216 | 1.0 | DorsAttnB | | | DorsalAttention |
| Occipital_Mid_L | L | 162 | 0.7 | VisPeri | | | Visual |
| SupraMarginal_L | L | 105 | 0.5 | SomMotB | | | Sensorimotor |
| **Cluster 8 \| 930 voxels \| peak z = +1.96 \| Mid Cingulate / Posterior Cingulate (bilateral)** | | | | | | | |
| Cingulate_Mid_R | R | 371 | 1.6 | SalVentAttnB | | | Salience |
| Cingulate_Mid_L | L | 250 | 1.1 | SalVentAttnB | | | Salience |
| Cingulate_Post_L | L | 74 | 0.3 | DefaultA | | | DefaultMode |
| Precuneus_R | R | 66 | 0.3 | DefaultC | | | DefaultMode |
| **Cluster 11 \| 850 voxels \| peak z = +1.82 \| Precuneus (bilateral)** | | | | | | | |
| Precuneus_L | L | 437 | 1.9 | | DefaultC | DefaultMode | |
| Precuneus_R | R | 397 | 1.7 | | DefaultC | DefaultMode | |
| **Cluster 5 \| 748 voxels \| peak z = +1.81 \| Middle / Inferior Temporal Right** | | | | | | | |
| Temporal_Mid_R | R | 561 | 2.5 | | DefaultA | DefaultMode | |
| Temporal_Inf_R | R | 187 | 0.8 | | DefaultA | DefaultMode | |
| **Cluster 4 \| 555 voxels \| peak z = +1.54 \| Middle / Inferior Temporal Left** | | | | | | | |
| Temporal_Mid_L | L | 344 | 1.5 | | DefaultA | DefaultMode | |
| Temporal_Inf_L | L | 187 | 0.8 | | DefaultA | DefaultMode | |
| **Cluster 1 \| 367 voxels \| peak z = +1.58 \| Cerebellum Crus1/2 Left** | | | | | | | |
| Cerebellum_Crus1_L | L | 271 | 1.2 | | Subcortical | Cerebellum | |
| Cerebellum_Crus2_L | L | 76 | 0.3 | | Subcortical | Cerebellum | |
| **Cluster 2 \| 112 voxels \| peak z = +1.41 \| Cerebellum Crus1 Right** | | | | | | | |
| Cerebellum_Crus1_R | R | 78 | 0.3 | | Subcortical | Cerebellum | |

| **FP-DM_CAP_ CO-DEACTIVATION** | | | | | |
| --- | --- | --- | --- | --- | --- |
| **Region_AAL3** | **Hemi** | **Voxels** | **% Total** | **Yeo17_Network** | **Functional System** |
| **Cluster 1 \| 18,153 voxels \| peak z = −2.79 \| Visual Cortex (occipital + lingual + fusiform)** | | | | | |
| Occipital_Mid_L | L | 1936 | 6.8 | VisPeri | Visual |
| Calcarine_L | L | 1562 | 5.5 | VisCent | Visual |
| Lingual_R | R | 1553 | 5.5 | VisCent | Visual |
| Lingual_L | L | 1406 | 5.0 | VisCent | Visual |
| Occipital_Mid_R | R | 1235 | 4.4 | VisPeri | Visual |
| Calcarine_R | R | 1230 | 4.4 | VisCent | Visual |
| Cuneus_R | R | 1029 | 3.6 | VisPeri | Visual |
| Cuneus_L | L | 1000 | 3.5 | VisPeri | Visual |
| Occipital_Sup_L | L | 916 | 3.2 | DorsAttnB | DorsalAttention |
| Fusiform_R | R | 814 | 2.9 | VisPeri | Visual |
| Occipital_Sup_R | R | 802 | 2.8 | DorsAttnB | DorsalAttention |
| Occipital_Inf_R | R | 728 | 2.6 | VisPeri | Visual |
| Occipital_Inf_L | L | 651 | 2.3 | VisPeri | Visual |
| Fusiform_L | L | 641 | 2.3 | VisPeri | Visual |
| Temporal_Mid_R | R | 559 | 2.0 | DefaultA | DefaultMode |
| Temporal_Inf_R | R | 194 | 0.7 | DefaultA | DefaultMode |
| Temporal_Mid_L | L | 134 | 0.5 | DefaultA | DefaultMode |
| Cerebellum_6_L | L | 358 | 1.3 | Subcortical | Cerebellum |
| Cerebellum_6_R | R | 299 | 1.1 | Subcortical | Cerebellum |
| Cerebellum_4_5_R | R | 86 | 0.3 | Subcortical | Cerebellum |
| **Cluster 2 \| 5,550 voxels \| peak z = −1.74 \| Precentral / Postcentral / SMA Right** | | | | | |
| Postcentral_R | R | 1737 | 6.1 | SomMotA | Sensorimotor |
| Precentral_R | R | 942 | 3.3 | SomMotA | Sensorimotor |
| Rolandic_Oper_R | R | 724 | 2.6 | SomMotB | Sensorimotor |
| Temporal_Sup_R | R | 478 | 1.7 | TempPar | Temporoparietal |
| Supp_Motor_Area_R | R | 271 | 1.0 | SomMotB | Sensorimotor |
| Paracentral_Lobule_R | R | 190 | 0.7 | SomMotA | Sensorimotor |
| SupraMarginal_R | R | 172 | 0.6 | SomMotB | Sensorimotor |
| Insula_R | R | 91 | 0.3 | SalVentAttnA | Salience |
| **Cluster 3 \| 4,568 voxels \| peak z = −1.78 \| Precentral / Postcentral / SMA Left** | | | | | |
| Postcentral_L | L | 2173 | 7.7 | SomMotA | Sensorimotor |
| Temporal_Sup_L | L | 759 | 2.7 | TempPar | Temporoparietal |
| Precentral_L | L | 553 | 2.0 | SomMotA | Sensorimotor |
| Rolandic_Oper_L | L | 482 | 1.7 | SomMotB | Sensorimotor |
| SupraMarginal_L | L | 133 | 0.5 | SomMotB | Sensorimotor |
| Insula_L | L | 70 | 0.2 | SalVentAttnA | Salience |

**Supplementary Table S3. FP-DM_CAP_ (Frontoparietal–Default Mode) co-activation and de-activation pattern: Regional composition and functional network assignment.** Anatomical regions comprising the FP-DM_CAP_ co-activation pattern (z ≥ +1.0, cluster ≥ 50 voxels; **co-activation:** 22,735 total voxels; 11 clusters, dominated by Control network [48.9%] and Default Mode [24.5%]) and de-activation pattern (z ≤ −1.0, cluster ≥ 50 voxels; de-coactivation: 28,271 total voxels; 3 clusters, dominated by Visual [47.0%] and Somatomotor [27.3%] networks). Regions with > 60 voxels are reported. Region_AAL3: Automated Anatomical Labelling Atlas v3.1 nomenclature. Hemi: L, left; R, right. Voxels: 2 mm³ MNI space. % Total: proportion of all voxels in the respective pattern. Yeo17_Network: assignment to Yeo’s 17-network parcellation (Schaefer et al., 2018). Functional System: higher-level cognitive system label.

| **VS-SC_CAP_ COACTIVATION** | | | | | |
| --- | --- | --- | --- | --- | --- |
| **Region_AAL3** | **Hemi** | **Voxels** | **% Total** | **Yeo17_Network** | **Functional System** |
| **Cluster 1 \| 37,995 voxels \| peak z = +3.43 \| Visual / Parieto-occipital / Sensorimotor (B)** | | | | | |
| Occipital_Mid_L | L | 2602 | 6.8 | VisPeri | Visual |
| Postcentral_R | R | 2025 | 5.3 | SomMotA | Sensorimotor |
| Calcarine_L | L | 1995 | 5.2 | VisCent | Visual |
| Postcentral_L | L | 1982 | 5.2 | SomMotA | Sensorimotor |
| Lingual_R | R | 1798 | 4.7 | VisCent | Visual |
| Occipital_Mid_R | R | 1738 | 4.5 | VisPeri | Visual |
| Lingual_L | L | 1655 | 4.3 | VisCent | Visual |
| Parietal_Sup_L | L | 1532 | 4.0 | DorsAttnB | DorsalAttention |
| Parietal_Sup_R | R | 1523 | 4.0 | DorsAttnB | DorsalAttention |
| Calcarine_R | R | 1461 | 3.8 | VisCent | Visual |
| Precuneus_R | R | 1423 | 3.7 | DefaultC | DefaultMode |
| Precuneus_L | L | 1421 | 3.7 | DefaultC | DefaultMode |
| Cuneus_L | L | 1239 | 3.2 | VisPeri | Visual |
| Cuneus_R | R | 1216 | 3.2 | VisPeri | Visual |
| Occipital_Sup_L | L | 1097 | 2.9 | DorsAttnB | DorsalAttention |
| Occipital_Sup_R | R | 1093 | 2.8 | DorsAttnB | DorsalAttention |
| Fusiform_R | R | 1048 | 2.7 | VisPeri | Visual |
| Fusiform_L | L | 903 | 2.3 | VisPeri | Visual |
| Occipital_Inf_R | R | 894 | 2.3 | VisPeri | Visual |
| Occipital_Inf_L | L | 882 | 2.3 | VisPeri | Visual |
| Temporal_Mid_R | R | 872 | 2.3 | DefaultA | DefaultMode |
| Precentral_R | R | 779 | 2.0 | SomMotA | Sensorimotor |
| Temporal_Inf_R | R | 565 | 1.5 | DefaultA | DefaultMode |
| Precentral_L | L | 509 | 1.3 | SomMotA | Sensorimotor |
| Cerebellum_6_L | L | 497 | 1.3 | Subcortical | Cerebellum |
| Cerebellum_6_R | R | 487 | 1.3 | Subcortical | Cerebellum |
| Parietal_Inf_L | L | 446 | 1.2 | DorsAttnB | DorsalAttention |
| Temporal_Mid_L | L | 375 | 1.0 | DefaultA | DefaultMode |
| Paracentral_Lobule_R | R | 310 | 0.8 | SomMotA | Sensorimotor |
| Cerebellum_Crus1_L | L | 237 | 0.6 | Subcortical | Cerebellum |
| Paracentral_Lobule_L | L | 205 | 0.5 | SomMotA | Sensorimotor |
| Cerebellum_Crus1_R | R | 157 | 0.4 | Subcortical | Cerebellum |
| Temporal_Inf_L | L | 126 | 0.3 | DefaultA | DefaultMode |
| Cerebellum_4_5_R | R | 120 | 0.3 | Subcortical | Cerebellum |
| Parietal_Inf_R | R | 117 | 0.3 | DorsAttnB | DorsalAttention |
| Cerebellum_4_5_L | L | 90 | 0.2 | Subcortical | Cerebellum |
| **Cluster 2 \| 234 voxels \| peak z = +1.29 \| Superior Temporal Gyrus / Heschl Left (auditory)** | | | | | |
| Temporal_Sup_L | L | 150 | 0.4 | TempPar | Temporoparietal |
| **Cluster 3 \| 220 voxels \| peak z = +1.25 \| Superior Temporal Gyrus / Heschl Right (auditory)** | | | | | |
| Temporal_Sup_R | R | 125 | 0.3 | TempPar | Temporoparietal |

**Supplementary Table S4. VS-SC_CAP_ (Visual–Somatomotor) co-activation pattern:** Regional composition and functional network assignment. Anatomical regions comprising the VS-SC_CAP_ co-activation pattern (z ≥ +1.0, cluster ≥ 50 voxels; 38,449 total voxels; 3 clusters, dominated by Visual [40.4%], Dorsal Attention [22.0%], and Somatomotor [13.3%] networks). No de-activation clusters (z ≤ −1.0, ≥ 50 voxels) were identified; VS-SC_CAP_ is therefore a coactivation-only state. Regions with > 60 voxels are reported. Region_AAL3: Automated Anatomical Labelling Atlas v3.1 nomenclature. Hemi: L, left; R, right. Voxels: 2 mm³ MNI space. % Total: proportion of all VS-SC_CAP_ co-activation voxels. Yeo17_Network: assignment to Yeo’s 17-network parcellation (Schaefer et al., 2018). Functional System: higher-level cognitive system label.

|  |  | **AFF (*N*= 228)** | | | |  | **CTR (*N*= 112)** | | | |  | **Mann–Whitney AFF vs CTR** | | | |
| --- | --- | --- | --- | --- | --- | --- | --- | --- | --- | --- | --- | --- | --- | --- | --- |
| **Variable** |  | **Mean** | **SD** | **Median** | **IQR** |  | **Mean** | **SD** | **Median** | **IQR** |  | **MW W** | ***p*** | **rb** | ***q*** |
| **CAP DWELL TIME (seconds)** | | | | | | | | | | | | | | | |
| AT-SM |  | 4.72 | 0.86 | 4.60 | 1.21 |  | 4.81 | 0.95 | 4.71 | 1.11 |  | 12388 | 0.66 | 0.03 | 0.87 |
| DM |  | 4.67 | 1.00 | 4.60 | 1.13 |  | 4.67 | 0.95 | 4.60 | 1.16 |  | 12737 | 0.97 | 0.00 | 0.97 |
| FP-DM |  | 4.43 | 0.82 | 4.31 | 1.07 |  | 4.64 | 0.85 | 4.52 | 1.26 |  | 10830 | 0.02 | 0.15 | 0.09 |
| VS-SC |  | 4.22 | 0.91 | 4.11 | 1.14 |  | 4.13 | 0.84 | 4.11 | 1.23 |  | 13322 | 0.52 | -0.04 | 0.87 |
| **CAP ENTRIES (count)** | | | | | | | | | | | | | | | |
| AT-SM |  | 25.12 | 4.36 | 25.00 | 6.00 |  | 24.96 | 3.78 | 25.00 | 6.00 |  | 13058 | 0.73 | -0.02 | 0.88 |
| DM |  | 24.40 | 5.36 | 24.00 | 7.00 |  | 24.51 | 4.78 | 25.00 | 6.00 |  | 12639 | 0.88 | 0.01 | 0.88 |
| FP-DM |  | 24.67 | 3.94 | 25.00 | 5.00 |  | 24.95 | 3.83 | 25.00 | 6.00 |  | 12402 | 0.67 | 0.03 | 0.88 |
| VS-SC |  | 25.00 | 6.67 | 25.00 | 9.00 |  | 25.80 | 6.20 | 26.00 | 8.25 |  | 12011 | 0.37 | 0.06 | 0.88 |
| **SYMPTOMS** | | | | | | | | | | | | | | | |
| IDS |  | 28.22 | 11.29 | 29.00 | 14.00 |  | 6.23 | 5.51 | 5.00 | 7.00 |  | 24280 | < .001 | -0.91 | < .001 |
| BAI |  | 14.37 | 9.31 | 13.00 | 12.00 |  | 2.48 | 3.92 | 1.00 | 2.20 |  | 23302 | < .001 | -0.83 | < .001 |
| FQ |  | 1.95 | 1.28 | 2.00 | 2.00 |  | 0.46 | 0.70 | 0.00 | 1.00 |  | 17668 | < .001 | -0.71 | < .001 |

**Table S5. Cross-sectional descriptive statistics and group comparisons of CAP dwell time (DUR), CAP entries (ENT), and symptom scores at baseline for affective patients (AFF: MDD and anxiety disorder) and healthy controls (CTR).** Values are derived from the first available wave per subject (earliest of W1/W3/W6). CAP metrics are derived from the K=4 CAP solution applied to NESDA resting-state fMRI data. DUR = mean dwell time per CAP state (seconds); ENT = number of entries into each CAP state per session. Symptom scores: IDS = Inventory of Depressive Symptomatology total score; BAI = Beck Anxiety Inventory total score; FQ = Fear Questionnaire total severity score (fearqtsc). Group comparisons were performed using two-sided Mann–Whitney U tests. “rb” = rank-biserial correlation (effect size; positive values indicate higher values in CTR, negative values indicate higher values in AFF). *q* = Benjamini–Hochberg false discovery rate-corrected p-value, applied separately within each metric block (4 CAP tests per block). No group difference in DUR or ENT survived FDR correction. Symptom scores differed markedly between groups across all three measures (all *q* < .001), confirming expected clinical divergence.

| **symptom** | **rho_AFF_** | **rho_CTR_** | **Δrho** | **CI_LOW_** | **CI_HIGH_** | **Z stat** | ***p*** | ***q*** |
| --- | --- | --- | --- | --- | --- | --- | --- | --- |
| **AT-SM_CAP_** | | | | | | | | |
| BAI | 0 | -0.1 | 0.1 | -0.15 | 0.33 | 0.78 | 0.43 | 0.58 |
| FQ | -0.06 | -0.11 | 0.05 | -0.2 | 0.29 | 0.35 | 0.72 | 0.94 |
| IDS | 0.09 | -0.29 | 0.36 | 0.13 | 0.56 | 2.99 | <0.001 | 0.01 |
| **DM_CAP_** | | | | | | | | |
| BAI | -0.07 | -0.08 | 0.02 | -0.23 | 0.26 | 0.13 | 0.89 | 0.89 |
| FQ | -0.06 | -0.07 | 0.01 | -0.24 | 0.25 | 0.07 | 0.94 | 0.94 |
| IDS | 0 | -0.23 | 0.24 | -0.01 | 0.45 | 1.89 | 0.06 | 0.12 |
| **FP-DM_CAP_** | | | | | | | | |
| BAI | -0.05 | -0.18 | 0.13 | -0.12 | 0.36 | 1.04 | 0.3 | 0.58 |
| FQ | -0.13 | -0.01 | -0.12 | -0.35 | 0.13 | -0.91 | 0.36 | 0.94 |
| IDS | -0.02 | -0.16 | 0.14 | -0.11 | 0.37 | 1.11 | 0.27 | 0.27 |
| **VS-SC_CAP_** | | | | | | | | |
| BAI | 0.01 | -0.21 | 0.22 | -0.03 | 0.44 | 1.76 | 0.08 | 0.31 |
| FQ | 0.04 | 0.02 | 0.02 | -0.23 | 0.26 | 0.13 | 0.9 | 0.94 |
| IDS | -0.01 | -0.16 | 0.15 | -0.1 | 0.38 | 1.19 | 0.23 | 0.27 |

**Supplementary Table S6A. Cross-sectional analysis. Group differences in CAP dwell time–symptom associations at baseline.** For each CAP state (AT-SM_CAP_–VS-SC_CAP_) and three symptom dimensions — depression (IDS), anxiety (BAI), and fear (FQ) — the table reports the partial Spearman correlation in affective patients (rho_AFF,_ n = 225) and controls (rho_CTR_, n = 92), the between-group difference (Δrho = rho_AFF_ − rho_CTR_), its 95% confidence interval (CI_LOW_, CI_HIGH_), Fisher z-statistic, uncorrected p-value, and FDR-corrected q-value (Benjamini–Hochberg, applied across 4 CAPs within each symptom). Partial correlations are adjusted for sex, age, scanning site, medication. The AT-SM_CAP_–IDS contrast was the only significant between-group difference (Δrho = 0.36 [0.13, 0.56], q = 0.01), supporting Figure 3C. No significant between-group differences were observed for anxiety or fear, or for any other CAP metric.

| **symptom** | **rho_AFF_** | **rho_CTR_** | **Δrho** | **CI_LOW_** | **CI_HIGH_** | **Z stat** | ***p*** | ***q*** |
| --- | --- | --- | --- | --- | --- | --- | --- | --- |
| **AT-SM_CAP_** |  |  |  |  |  |  |  |  |
| BAI | 0.03 | 0.03 | 0.00 | -0.25 | 0.25 | 0.02 | 0.99 | 0.99 |
| FQ | 0.11 | 0.04 | 0.07 | -0.19 | 0.31 | 0.52 | 0.60 | 0.81 |
| IDS | 0.07 | 0.16 | -0.09 | -0.33 | 0.16 | -0.72 | 0.47 | 0.63 |
| **DM_CAP_** |  |  |  |  |  |  |  |  |
| BAI | -0.04 | 0.10 | -0.14 | -0.37 | 0.12 | -1.05 | 0.29 | 0.99 |
| FQ | 0.04 | -0.09 | 0.13 | -0.12 | 0.37 | 1.03 | 0.30 | 0.81 |
| IDS | -0.02 | 0.01 | -0.03 | -0.28 | 0.22 | -0.23 | 0.82 | 0.82 |
| **FP-DM_CAP_** |  |  |  |  |  |  |  |  |
| BAI | 0.04 | -0.02 | 0.05 | -0.20 | 0.30 | 0.39 | 0.70 | 0.99 |
| FQ | 0.03 | 0.14 | -0.10 | -0.34 | 0.15 | -0.79 | 0.43 | 0.81 |
| IDS | 0.06 | 0.21 | -0.16 | -0.39 | 0.10 | -1.23 | 0.22 | 0.44 |
| **VS-SCCAP** |  |  |  |  |  |  |  |  |
| BAI | -0.05 | -0.01 | -0.04 | -0.29 | 0.21 | -0.31 | 0.75 | 0.99 |
| FQ | 0.01 | 0.01 | 0.00 | -0.25 | 0.25 | -0.03 | 0.98 | 0.98 |
| IDS | -0.07 | 0.18 | -0.24 | -0.47 | 0.01 | -1.92 | 0.06 | 0.22 |

**Supplementary Table S6B. Cross-sectional analysis. Group differences in CAP entries–symptom associations at baseline.** For each CAP state (AT-SM_CAP–VS-SC_CAP) and three symptom dimensions — depression (IDS), anxiety (BAI), and fear (FQ) — the table reports the partial Spearman correlation between CAP entries and symptom severity in affective patients (rho_AFF, n = 225) and controls (rho_CTR, n = 92), the between-group difference (Δrho = rho_AFF − rho_CTR), its 95% confidence interval (CI_LOW, CI_HIGH), Fisher z-statistic, uncorrected *p*-value, and FDR-corrected *q*-value (Benjamini–Hochberg, applied across 4 CAPs within each symptom). Partial correlations are adjusted for sex, age, scanning site, medication. No between-group difference in CAP entries–symptom coupling reached significance for any CAP or symptom (all *q* ≥ 0.22), consistent with the entries metric showing weaker cross-sectional signal than dwell time (Table S6A).

| **VARIABLE** | ***F*** | **df1** | **df2** | ***p*** | **η²p** | ***q*** |
| --- | --- | --- | --- | --- | --- | --- |
| **CAP DWELL TIME** |  |  |  |  |  |  |
| AT-SM | 0.66 | 2 | 478.00 | 0.52 | 0.00 | 0.86 |
| DM | 0.15 | 2 | 510.50 | 0.86 | 0.00 | 0.86 |
| FP-DM | 1.47 | 2 | 467.30 | 0.23 | 0.01 | 0.86 |
| VS-SC | 0.41 | 2 | 510.50 | 0.66 | 0.00 | 0.86 |
| **CAP ENTRIES** |  |  |  |  |  |  |
| AT-SM | 0.72 | 2 | 499.00 | 0.49 | 0.00 | 0.83 |
| DM | 0.21 | 2 | 499.60 | 0.81 | 0.00 | 0.83 |
| FP-DM | 1.47 | 2 | 495.50 | 0.23 | 0.01 | 0.83 |
| VS-SC | 0.18 | 2 | 510.50 | 0.83 | 0.00 | 0.83 |
| **SYMPTOMS** |  |  |  |  |  |  |
| IDS | 7.84 | 2 | 397.20 | <.001 | 0.04 | <.001 |
| BAI | 5.80 | 2 | 359.40 | <.01 | 0.03 | <.01 |
| FQ | 1.52 | 2 | 315.10 | 0.22 | 0.01 | 0.22 |

**Supplementary Table S7a. Group × session interactions in CAP dynamics and symptom severity.** **Omnibus testing.** Group (AFF, CTR) × session (Y0, Y2, Y9) interaction tests from linear mixed models with random intercepts for participant (model: *z_y ~ group*_lmm *× session + covariates + (1 | pident)),* estimated with REML and Kenward-Roger approximation for degrees of freedom, adjusted for age, sex, and scanning site. The interaction term tests whether the shape of change over time differs between groups, distinct from the within-group session effect (Table S8, S9) and the simple between-group contrasts at each wave (Table S10, S11). F, df1, df2, and p refer to the omnibus interaction test; η²p is partial eta-squared, computed as *(F × df1) / (F × df1 + df2)*. p-values (*q*) were corrected for multiple comparisons using the Benjamini–Hochberg false discovery rate (FDR) method, applied separately within CAP dwell time, CAP entries, and symptom families (4 CAPs per metric x 3 symptoms). CAP dwell time = dwell duration (DUR metric); CAP entries = state entry count (ENT metric); IDS = Inventory of Depressive Symptomatology; BAI = Beck Anxiety Inventory; FQ = Fear Questionnaire. No CAP metric showed a significant group × time interaction (all *q* ≥ 0.83). Depression and anxiety trajectories differed significantly by group (IDS, BAI: *q* < 0.01); fear trajectories did not (*q* = 0.22).

| **Session (years)** | **β (AFF−CTR)** | ***d*** | **SE** | **df** | ***t*** | ***p*** |
| --- | --- | --- | --- | --- | --- | --- |
| **IDS** |  |  |  |  |  |  |
| Y0 | 1.54 | 2.20 | 0.10 | 469.53 | 15.25 | <.001 |
| Y2 | 1.02 | 1.45 | 0.11 | 414.61 | 9.69 | <.001 |
| Y9 | 1.15 | 1.65 | 0.11 | 498.34 | 10.81 | <.001 |
| **BAI** |  |  |  |  |  |  |
| Y0 | 1.25 | 1.49 | 0.11 | 439.61 | 10.89 | <.001 |
| Y2 | 0.79 | 0.94 | 0.12 | 369.19 | 6.71 | <.001 |
| Y9 | 0.80 | 0.96 | 0.12 | 473.91 | 6.60 | <.001 |
| **FQ** |  |  |  |  |  |  |
| Y0 | 0.86 | 0.98 | 0.11 | 387.07 | 7.72 | <.001 |
| Y2 | 0.61 | 0.70 | 0.11 | 324.10 | 5.43 | <.001 |
| Y9 | 0.78 | 0.88 | 0.12 | 403.38 | 6.24 | <.001 |

**Supplementary Table S7b. Between-group comparisons (AFF vs. CTR) at each wave of symptoms severity scores.** Between-group contrasts from linear mixed models (LMMs) with group (AFF, CTR) × session (Y0, Y2, Y9) interaction and random intercepts for participant (model: z_y ~ group_lmm × wave + covariates + (1 | pident)), estimated with REML and Kenward-Roger approximation for degrees of freedom, adjusted for age, sex, and scanning site. Comparisons are presented at baseline (Y0/w1), 2-year follow-up (Y2/w3), and 9-year follow-up (Y9/w6). For each wave, the table reports the contrast estimate (β), Cohen's *d* effect size (computed as estimate/σ_total, where σ_total = √(variance_random_intercept + variance_residual)), standard error (SE), degrees of freedom (df), *t*-statistic, and *p*-value. Depression = Inventory of Depressive Symptomatology (IDS); Anxiety = Beck Anxiety Inventory (BAI); Fear = Fear Questionnaire (FQ). AFF vs. CTR differed significantly at every wave for all three symptom measures (all *p* < .001), reflecting the large and sustained baseline symptom-severity gap between groups. Corresponding omnibus group × time interaction tests are reported in Table S7.

| **CAP** | **Session** | **β (AFF vs. CTR)** | ***d*** | **SE** | **df** | ***t*** | ***p*** |
| --- | --- | --- | --- | --- | --- | --- | --- |
| **DWELL TIME** | |  |  |  |  |  |  |
| AT-SM | Y0 | −0.03 | −0.03 | 0.15 | 526.00 | −0.17 | 0.86 |
| AT-SM | Y2 | 0.07 | 0.07 | 0.16 | 514.72 | 0.44 | 0.66 |
| AT-SM | Y9 | 0.22 | 0.22 | 0.15 | 529.99 | 1.40 | 0.16 |
| DM | Y0 | 0.04 | 0.04 | 0.15 | 532.98 | 0.23 | 0.82 |
| DM | Y2 | 0.12 | 0.12 | 0.16 | 532.97 | 0.76 | 0.45 |
| DM | Y9 | 0.15 | 0.15 | 0.16 | 532.98 | 0.96 | 0.34 |
| FP-DM | Y0 | −0.37 | −0.38 | 0.15 | 521.79 | −2.50 | 0.01 |
| FP-DM | Y2 | −0.15 | −0.15 | 0.16 | 503.70 | −0.94 | 0.35 |
| FP-DM | Y9 | −0.02 | −0.02 | 0.15 | 528.06 | −0.10 | 0.92 |
| VS-SC | Y0 | −0.03 | −0.03 | 0.15 | 532.98 | −0.20 | 0.84 |
| VS-SC | Y2 | −0.10 | −0.11 | 0.16 | 532.97 | −0.64 | 0.52 |
| VS-SC | Y9 | 0.10 | 0.10 | 0.16 | 532.98 | 0.62 | 0.53 |
| **ENTRIES** | |  |  |  |  |  |  |
| AT-SM | Y0 | 0.11 | 0.12 | 0.15 | 531.77 | 0.74 | 0.46 |
| AT-SM | Y2 | −0.15 | −0.15 | 0.16 | 530.38 | −0.94 | 0.35 |
| AT-SM | Y9 | −0.03 | −0.03 | 0.16 | 532.60 | −0.19 | 0.85 |
| DM | Y0 | 0.13 | 0.13 | 0.15 | 531.87 | 0.83 | 0.41 |
| DM | Y2 | 0.07 | 0.07 | 0.16 | 530.66 | 0.43 | 0.67 |
| DM | Y9 | −0.01 | −0.01 | 0.16 | 532.64 | −0.09 | 0.93 |
| FP-DM | Y0 | −0.15 | −0.15 | 0.15 | 531.08 | −0.94 | 0.35 |
| FP-DM | Y2 | 0.18 | 0.18 | 0.16 | 528.55 | 1.11 | 0.27 |
| FP-DM | Y9 | −0.16 | −0.16 | 0.16 | 532.30 | −1.01 | 0.31 |
| VS-SC | Y0 | −0.09 | −0.09 | 0.16 | 532.98 | −0.57 | 0.57 |
| VS-SC | Y2 | −0.18 | −0.18 | 0.16 | 532.97 | −1.09 | 0.28 |
| VS-SC | Y9 | −0.22 | −0.22 | 0.16 | 532.98 | −1.37 | 0.17 |

**Supplementary Table S7c. Between-group comparisons (AFF vs. CTR) at each wave of CAP metrics.** Between-group contrasts from linear mixed models (LMMs) with group (AFF, CTR) × time (Y0, Y2, Y9) interaction and random intercepts for participant (model: *z_y ~ group_lmm × wave + covariates + (1 | pident)),* estimated with REML and Kenward-Roger approximation for degrees of freedom, adjusted for age, sex, and scanning site. Comparisons are presented at baseline (Y0), 2-year follow-up (Y2), and 9-year follow-up (Y9). For each wave, the table reports the contrast estimate (β), Cohen's d effect size (computed as estimate/σ_total, where σ_total = √(variance_random_intercept + variance_residual)), standard error (*SE*), degrees of freedom (*df*), t-statistic, and p-value. CAP dwell time = dwell duration (DUR metric); CAP entries = state entry count (ENT metric). No between-group contrast survived correction for multiple comparisons (all corresponding q > 0.05; Table S7a); the single nominally significant cell, FP-DM_CAP_ dwell time at Y0 (p = 0.01), did not remain significant after FDR correction and should be interpreted alongside the non-significant omnibus group × time interaction for FP-DM_CAP_ (Table S7a).

|  | **Y0 → Y2  (n = 82)** | | | **Y0 → Y9  (n = 49)** | | | **Y2 → Y9  (n = 39)** | | |
| --- | --- | --- | --- | --- | --- | --- | --- | --- | --- |
|  | **β [95% CI]** | ***d*** | ***q*** | **β [95% CI]** | ***d*** | ***q*** | **β [95% CI]** | ***d*** | ***q*** |
| **CAP DWELL TIME** | | | | | | | | | |
| AT-SM | 0.03 [−0.27, +0.33] | 0.03 | 0.85 | 0.85 [+0.49, +1.22] | 0.91 | <0.001*** | 0.83 [0.42, 1.23] | 0.88 | <0.001*** |
| DM | 0.09 [−0.25, +0.43] | 0.09 | 0.58 | 0.66 [+0.25, +1.06] | 0.66 | <0.01** | 0.56 [0.11, 1.01] | 0.57 | <0.05* |
| FP-DM | −0.22 [−0.52, +0.09] | −0.23 | 0.16 | +0.37 [+0.00, +0.75] | 0.39 | 0.073† | 0.59 [0.18, 1.01] | 0.62 | <0.05* |
| VS-SC | −0.14 [−0.48, +0.20] | −0.14 | 0.85 | −0.01 [−0.42, +0.41] | −0.01 | 0.980 | 0.14 [−0.33, 0.60] | 0.13 | 0.847 |
| **CAP ENTRIES** | | | | | | | | | |
| AT-SM | +0.05 [−0.28, +0.38] | +0.05 | 0.762 | −0.25 [−0.65, +0.15] | −0.25 | 0.328 | −0.30 [−0.74, +0.14] | −0.30 | 0.328 |
| DM | −0.13 [−0.47, +0.22] | −0.12 | 0.569 | −0.26 [−0.67, +0.15] | −0.25 | 0.569 | −0.13 [−0.59, +0.33] | −0.13 | 0.569 |
| FP-DM | +0.08 [−0.25, +0.41] | +0.08 | 0.649 | −0.39 [−0.79, +0.01] | −0.39 | 0.086† | −0.46 [−0.91, −0.02] | −0.47 | 0.086† |
| VS-SC | −0.01 [−0.36, +0.34] | −0.01 | 0.968 | −0.16 [−0.58, +0.27] | −0.15 | 0.798 | −0.15 [−0.61, +0.32] | −0.15 | 0.798 |
| **SYMPTOMS** | | | | | | | | | |
| DEPRESSION | −0.33 [−0.52, −0.14] | −0.46 | <0.01** | −0.35 [−0.58, −0.12] | −0.49 | <0.01** | −0.02 [−0.27, 0.24] | −0.03 | 0.884 |
| ANXIETY | −0.25 [−0.45, −0.05] | −0.34 | <0.05* | −0.23 [−0.47, 0.01] | −0.32 | 0.092† | +0.02 [−0.25, 0.29] | 0.03 | 0.884 |
| FEAR | −0.18 [−0.37, +0.02] | −0.21 | 0.221 | −0.13 [−0.38, +0.11] | −0.16 | 0.412 | +0.04 [−0.22, 0.31] | 0.05 | 0.746 |

**Supplementary Table S8. Within-group longitudinal change. AFF group.**  Linear mixed model with random intercept per subject: z-outcome ~ group × wave + (1 | pident); Kenward-Roger degrees of freedom. β = estimated marginal mean difference (z-score units); 95% CI model-based. d = Cohen's d (β / σtotal; σtotal = √(τ² + σ²) from model). *q*: Benjamini-Hochberg applied per variable within its three wave contrasts (family size = 3). CAP dwell time = dwell time; CAP entries = state entry count. Symptoms shared across metrics (identical analytic sample). n = subjects with valid data at both waves of each contrast. *** *q* < 0.001   ** < 0.01   * < 0.05   † < 0.10 trend   (blank) ns.

|  | ***Y0 → Y2  (n = 34)*** | | | ***Y0 → Y9  (n = 18)*** | | | ***Y2 → Y9  (n = 21)*** | | |
| --- | --- | --- | --- | --- | --- | --- | --- | --- | --- |
|  | **β [95% CI]** | ***d*** | ***q*** | **β [95% CI]** | ***d*** | ***q*** | **β [95% CI]** | ***d*** | ***q*** |
| **CAP DWELL TIME** | | | | | | | | | |
| AT-SM | −0.18 [−0.53, +0.16] | −0.19 | 0.299 | +0.46 [+0.08, +0.84] | +0.49 | <0.05* | +0.64 [+0.33, +0.96] | +0.69 | <0.001*** |
| DM | +0.04 [−0.35, +0.42] | +0.04 | 0.844 | +0.63 [+0.21, +1.05] | +0.64 | <0.01** | +0.59 [+0.24, +0.94] | +0.60 | <0.01** |
| FP-DM | −0.27 [−0.61, +0.08] | −0.28 | 0.134 | +0.37 [−0.01, +0.75] | +0.39 | *0.087†* | +0.64 [+0.32, +0.96] | +0.66 | <0.001*** |
| VS-SC | +0.11 [−0.28, +0.50] | +0.11 | 0.800 | +0.07 [−0.36, +0.49] | +0.07 | 0.800 | −0.05 [−0.40, +0.31] | −0.05 | 0.800 |
| **CAP ENTRIES** | | | | | | | | | |
| AT-SM | +0.05 [−0.32, +0.43] | +0.05 | 0.775 | −0.27 [−0.68, +0.14] | −0.28 | 0.282 | −0.33 [−0.67, +0.01] | −0.33 | 0.180 |
| DM | −0.18 [−0.57, +0.21] | −0.17 | 0.365 | −0.34 [−0.77, +0.08] | −0.33 | 0.339 | −0.16 [−0.52, +0.19] | −0.16 | 0.365 |
| FP-DM | −0.15 [−0.52, +0.23] | −0.15 | 0.440 | −0.48 [−0.89, −0.07] | −0.49 | *0.067†* | −0.33 [−0.67, +0.01] | −0.34 | *0.088†* |
| VS-SC | +0.06 [−0.33, +0.46] | +0.06 | 0.757 | −0.07 [−0.51, +0.36] | −0.07 | 0.757 | −0.14 [−0.50, +0.23] | −0.13 | 0.757 |
| **SYMPTOMS** | | | | | | | | | |
| DEPRESSION | +0.14 [−0.08, +0.35] | +0.19 | 0.314 | +0.16 [−0.08, +0.40] | +0.23 | 0.314 | +0.02 [−0.17, +0.22] | +0.03 | 0.814 |
| ANXIETY | +0.11 [−0.11, +0.34] | +0.16 | 0.471 | +0.15 [−0.10, +0.39] | +0.20 | 0.471 | +0.03 [−0.18, +0.24] | +0.04 | 0.767 |
| FEAR | +0.09 [−0.13, +0.31] | +0.11 | 0.621 | −0.05 [−0.30, +0.20] | −0.06 | 0.670 | −0.15 [−0.35, +0.06] | −0.17 | 0.496 |

**Supplementary Table S9. Within-group longitudinal change. CTR group.** Linear mixed model with random intercept per subject: *z-outcome ~ group × wave + (1 | pident);* Kenward-Roger degrees of freedom. β = estimated marginal mean difference (z-score units); 95% CI model-based. d = Cohen's d (β / σtotal; σtotal = √(τ² + σ²) from model). *q*: Benjamini-Hochberg applied per variable within its three wave contrasts (family size = 3). CAP DWELL TIME = dwell duration (DUR metric); CAP ENTRIES = state entry count (ENT metric). Symptoms shared across metrics (identical analytic sample). n = subjects with valid data at both waves of each contrast. *** *q* < 0.001   ** < 0.01   * < 0.05   † < 0.10 trend   (blank) ns.

| **CAP** | **Symptom** | **β s_w**  **CTR** | **β s_w**  **AFF** | **Δβ**  **(AFF−CTR)** | **p**  **(s_w × group)** | **LRT p**  **(M0 vs M1)** |
| --- | --- | --- | --- | --- | --- | --- |
| **CAP DWELL TIME** | | | | | | |
| AT-SM | IDS | -13.31 | -5.75 | 7.56 | 0.67 | 0.76 |
| DM | IDS | -37.73 | -17.69 | 20.04 | 0.25 | 0.37 |
| FP-DM | IDS | -3.47 | 9.05 | 12.52 | 0.47 | 0.50 |
| VS-SC | IDS | -23.31 | 11.29 | 34.61 | 0.05 | 0.25 |
| AT-SM | BAI | -7.51 | -1.09 | 6.42 | 0.70 | 0.92 |
| DM | BAI | -23.46 | -8.48 | 14.98 | 0.37 | 0.76 |
| FP-DM | BAI | -10.89 | -1.29 | 9.61 | 0.56 | 0.49 |
| VS-SC | BAI | -14.05 | 1.98 | 16.03 | 0.33 | 0.76 |
| AT-SM | FQ | -10.88 | 4.38 | 15.25 | 0.32 | 0.62 |
| DM | FQ | -14.43 | 1.88 | 16.31 | 0.30 | 0.69 |
| FP-DM | FQ | -3.81 | 2.24 | 6.05 | 0.69 | 0.46 |
| VS-SC | FQ | 16.31 | 9.52 | -6.79 | 0.66 | 0.58 |
| **CAP ENTRIES** | | | | | | |
| AT-SM | IDS | 26.57 | 6.13 | -20.44 | 0.24 | 0.18 |
| DM | IDS | 26.68 | 3.45 | -23.23 | 0.18 | 0.54 |
| FP-DM | IDS | -2.56 | 11.91 | 14.47 | 0.41 | 0.62 |
| VS-SC | IDS | 13.22 | 7.87 | -5.36 | 0.76 | 0.80 |
| AT-SM | BAI | 11.39 | 15.56 | 4.16 | 0.80 | 0.40 |
| DM | BAI | 35.98 | 2.80 | -33.18 | 0.04 | 0.26 |
| FP-DM | BAI | 6.10 | 1.23 | -4.87 | 0.77 | 0.99 |
| VS-SC | BAI | -1.49 | 5.91 | 7.40 | 0.66 | 0.65 |
| AT-SM | FQ | -3.05 | 6.01 | 9.06 | 0.56 | 0.49 |
| DM | FQ | 1.37 | -2.30 | -3.66 | 0.81 | 0.46 |
| FP-DM | FQ | 0.72 | 0.96 | 0.23 | 0.99 | 0.94 |
| VS-SC | FQ | -10.36 | -7.57 | 2.79 | 0.86 | 0.53 |

**Supplementary Table S10a. Diagnostic within-person coupling test (AFF vs. CTR) for the pooled group-interaction model.** For each CAP metric (dwell time, DUR; entries, ENT) × symptom (IDS, BAI, FQ) cell, a reduced model without a group term (M0: *rank(CAP) ~ s_b + s_w + covariates + (1|pident)*) was compared by likelihood-ratio test (LRT p, M0 vs M1) against the full group-interaction model used for inference in the manuscript (M1: *rank(CAP) ~ (s_b + s_w) × group + covariates + (1|pident);* covariates: age, sex, scanning site, education level, drug use). *β s_w* CTR and *β s_w* AFF are the within-person coupling coefficients implied for each group under M1; Δβ (AFF−CTR) is the *s_w × group interaction coefficient* (i.e., the AFF-vs-CTR difference in coupling) with its p-value. This is a diagnostic cross-check of the group-interaction result reported in the main analysis, not an independent test.

| **CAP** | **Symptom** | **Levene p**  **(res. var.)** | **AIC**  **M1** | **AIC**  **M1 (var. by grp)** | **ΔAIC** | **LRT p**  **(var. by grp)** | **Singular**  **fit (M1)** | **Shapiro p**  **AFF** | **Shapiro p**  **CTR** |
| --- | --- | --- | --- | --- | --- | --- | --- | --- | --- |
| **CAP DWELL TIME (DUR)** | | | | | | | | | |
| **AT-SM** | IDS | 0.48 | 6956.2 | 6957.9 | 1.70 | 0.56 | No | <0.001 | <0.001 |
| **DM** | IDS | 0.43 | 6952.4 | 6954.4 | 2.00 | 0.88 | Yes | <0.001 | <0.001 |
| **FP-DM** | IDS | 0.55 | 6944.6 | 6946.5 | 1.80 | 0.69 | No | <0.001 | <0.001 |
| **VS-SC** | IDS | 0.68 | 6954.3 | 6956.3 | 2.00 | 0.95 | No | <0.001 | <0.001 |
| **AT-SM** | BAI | 0.44 | 6957.5 | 6959.1 | 1.60 | 0.55 | No | <0.001 | <0.001 |
| **DM** | BAI | 0.43 | 6959.9 | 6961.8 | 2.00 | 0.84 | Yes | <0.001 | <0.001 |
| **FP-DM** | BAI | 0.55 | 6944.3 | 6946.1 | 1.80 | 0.64 | No | <0.001 | <0.001 |
| **VS-SC** | BAI | 0.71 | 6958.6 | 6960.5 | 2.00 | 0.91 | No | <0.001 | <0.001 |
| **AT-SM** | FQ | 0.35 | 6682.6 | 6684.1 | 1.50 | 0.48 | No | <0.001 | <0.001 |
| **DM** | FQ | 0.33 | 6694.4 | 6696.3 | 1.90 | 0.76 | Yes | <0.001 | <0.001 |
| **FP-DM** | FQ | 0.46 | 6667.1 | 6668.7 | 1.70 | 0.56 | No | <0.001 | <0.001 |
| **VS-SC** | FQ | 0.77 | 6680.6 | 6682.6 | 2.00 | 0.83 | No | <0.001 | <0.001 |
| **CAP ENTRIES (ENT)** | | | | | | | | | |
| **AT-SM** | IDS | 0.02 | 6943.6 | 6942.9 | -0.70 | 0.10 | No | <0.001 | <0.001 |
| **DM** | IDS | 0.23 | 6956.3 | 6957.9 | 1.70 | 0.56 | No | <0.001 | <0.001 |
| **FP-DM** | IDS | 0.73 | 6955.8 | 6957.8 | 2.00 | 0.86 | No | <0.001 | <0.001 |
| **VS-SC** | IDS | 0.93 | 6965.2 | 6967.1 | 2.00 | 0.84 | No | <0.001 | <0.001 |
| **AT-SM** | BAI | 0.04 | 6943.4 | 6943.2 | -0.20 | 0.14 | No | <0.001 | <0.001 |
| **DM** | BAI | 0.11 | 6955.1 | 6956.3 | 1.30 | 0.40 | No | <0.001 | <0.001 |
| **FP-DM** | BAI | 0.78 | 6958.7 | 6960.7 | 2.00 | 0.89 | No | <0.001 | <0.001 |
| **VS-SC** | BAI | 0.79 | 6968.7 | 6970.7 | 1.90 | 0.81 | No | <0.001 | <0.001 |
| **AT-SM** | FQ | 0.06 | 6676.5 | 6676.5 | 0.00 | 0.15 | No | <0.001 | <0.001 |
| **DM** | FQ | 0.19 | 6688.0 | 6689.8 | 1.80 | 0.65 | No | <0.001 | <0.001 |
| **FP-DM** | FQ | 0.76 | 6685.5 | 6687.4 | 2.00 | 0.92 | No | <0.001 | <0.001 |
| **VS-SC** | FQ | 0.79 | 6694.7 | 6696.7 | 2.00 | 0.83 | No | <0.001 | <0.001 |

**Supplementary Table S10b. Model-assumption diagnostics for the pooled group-interaction model.** Diagnostics testing whether AFF and CTR can be validly combined into a single pooled model (M1, as specified in Table S10a) rather than requiring group-specific structure. Levene’s test assesses the equal-residual-variance assumption of M1 across groups. AIC (M1) and AIC (M1, variance by group) compare the standard model to an otherwise identical model with group-specific residual variance (nlme, varIdent); ΔAIC is the difference (negative values favor the group-specific-variance model), and LRT p (var. by group) is the corresponding likelihood-ratio test. Singular fit flags convergence to a boundary solution for M1 in that cell. Shapiro–Wilk p-values assess residual normality within each group; because the outcome is rank-transformed, non-normal residuals are diagnostic rather than a modelling concern. Bold indicates p < 0.05. This table does not itself report an inferential result for the manuscript; it establishes whether the pooled group-interaction model (Supplementary Methods S6.4–S6.5) is an appropriate specification. **Interpretation of singular fits*.*** A singular fit indicates that the between-subject variance component of the random intercept was estimated at or near the lower boundary of zero (i.e., the data show little evidence of subject-level clustering in rank(CAP) beyond what the fixed effects and residual variance already explain. It does not mean that the model failed or that its fixed-effect estimates (including *β s_w* and the s*_w × group* interaction reported in Table S10a) are unreliable. All three singular fits in this table occur for the DM_CAP_ across all three symptoms (IDS, BAI, FQ) under the DUR metric, consistent with genuinely low between-subject variance in DM dwell time rather than a numerical artefact of any single fit. Because our primary longitudinal findings involve the FP-DM and VS-SC CAPs (main text §3.4.2), none of the singular fits reported here affect the manuscript’s reported coupling estimates. As a further check, fixed-effect estimates for the affected DM cells were compared against a random-intercept-free specification (ordinary least squares with cluster-robust standard errors by subject); point estimates for *β s_w* and the group interaction were consistent in sign and magnitude with the mixed-model estimates in Table S10a, indicating that the singular fits reflect a small random-intercept variance rather than a distortion of the coupling estimates themselves. **Levene-significant, AIC/LRT-nonsignificant cells*.*** Levene’s test was significant for the entries of AT-SM_CAP_ at two symptoms (IDS: *p* = 0.02; BAI: *p* = 0.04), flagging unequal residual variance between AFF and CTR in the pooled M1 model. However, the more decisive AIC/LRT comparison did not support modelling this heterogeneity explicitly: adding group-specific residual variance produced only a marginal AIC improvement in both cells (ΔAIC = −0.70 and −0.20, respectively) and the corresponding likelihood-ratio test was non-significant (*p* = 0.10 and *p* = 0.14). Levene’s test is more sensitive to small variance differences than the AIC/LRT comparison, which weighs whether accommodating that heterogeneity meaningfully improves model fit; the discrepancy here indicates a statistically detectable but practically minor departure from equal variance, not a violation serious enough to warrant a group-specific-variance specification. Neither cell corresponds to a CAP × metric combination reported in the manuscript’s primary findings (FP-DM and VS-SC dwell time; main text §3.4.2).

| **CAP** | **Symptom** | **β s_w**  **CTR** | **β s_w**  **AFF** | **Δβ**  **(AFF−CTR)** | **SE** | **t** | **df** | **p** | **η²p** | **q** |
| --- | --- | --- | --- | --- | --- | --- | --- | --- | --- | --- |
| **CAP DWELL TIME** | | | | | | | | | | |
| AT-SM | IDS | -13.18 | -5.72 | 7.46 | 17.69 | 0.42 | 522.3 | 0.673 | 0.0003 | 0.69 |
| DM | IDS | -37.73 | -17.69 | 20.04 | 17.50 | 1.15 | 524.0 | 0.253 | 0.0025 | 0.69 |
| FP-DM | IDS | -3.64 | 9.09 | 12.72 | 17.47 | 0.73 | 517.7 | 0.467 | 0.0010 | 0.69 |
| VS-SC | IDS | -23.34 | 11.32 | 34.67 | 17.57 | 1.97 | 501.8 | 0.049 | 0.0077 | 0.59 |
| AT-SM | BAI | -7.63 | -0.98 | 6.65 | 16.74 | 0.40 | 523.9 | 0.691 | 0.0003 | 0.69 |
| DM | BAI | -23.46 | -8.48 | 14.98 | 16.72 | 0.90 | 524.0 | 0.371 | 0.0015 | 0.69 |
| FP-DM | BAI | -11.07 | -1.26 | 9.81 | 16.48 | 0.59 | 510.5 | 0.552 | 0.0007 | 0.69 |
| VS-SC | BAI | -14.15 | 2.05 | 16.20 | 16.73 | 0.97 | 515.4 | 0.333 | 0.0018 | 0.69 |
| AT-SM | FQ | -10.95 | 4.41 | 15.36 | 15.56 | 0.99 | 468.3 | 0.324 | 0.0021 | 0.69 |
| DM | FQ | -14.43 | 1.88 | 16.31 | 15.88 | 1.03 | 503.0 | 0.305 | 0.0021 | 0.69 |
| FP-DM | FQ | -3.96 | 2.29 | 6.25 | 15.10 | 0.41 | 426.4 | 0.679 | 0.0004 | 0.69 |
| VS-SC | FQ | 16.49 | 9.45 | -7.04 | 15.65 | -0.45 | 495.6 | 0.653 | 0.0004 | 0.69 |
| **CAP ENTRIES** | | | | | | | | | | |
| AT-SM | IDS | 26.84 | 5.95 | -20.88 | 17.45 | -1.20 | 512.0 | 0.232 | 0.0028 | 0.92 |
| DM | IDS | 26.78 | 3.36 | -23.42 | 17.66 | -1.33 | 516.3 | 0.186 | 0.0034 | 0.92 |
| FP-DM | IDS | -2.50 | 11.87 | 14.37 | 17.66 | 0.81 | 518.0 | 0.416 | 0.0013 | 0.92 |
| VS-SC | IDS | 13.12 | 7.93 | -5.20 | 17.77 | -0.29 | 506.1 | 0.77 | 0.0002 | 0.92 |
| AT-SM | BAI | 11.77 | 15.35 | 3.58 | 16.52 | 0.22 | 520.5 | 0.829 | 0.0001 | 0.92 |
| DM | BAI | 36.20 | 2.63 | -33.57 | 16.70 | -2.01 | 523.1 | 0.045 | 0.0077 | 0.54 |
| FP-DM | BAI | 6.32 | 1.19 | -5.13 | 16.76 | -0.31 | 523.6 | 0.76 | 0.0002 | 0.92 |
| VS-SC | BAI | -1.56 | 5.96 | 7.51 | 16.89 | 0.45 | 517.6 | 0.657 | 0.0004 | 0.92 |
| AT-SM | FQ | -2.98 | 5.91 | 8.88 | 15.54 | 0.57 | 482.1 | 0.568 | 0.0007 | 0.92 |
| DM | FQ | 1.58 | -2.43 | -4.00 | 15.72 | -0.26 | 486.0 | 0.799 | 0.0001 | 0.92 |
| FP-DM | FQ | 0.94 | 0.86 | -0.08 | 15.63 | -0.01 | 477.5 | 0.996 | 0.0000 | 1.00 |
| VS-SC | FQ | -10.59 | -7.47 | 3.12 | 15.87 | 0.20 | 495.7 | 0.844 | 0.0001 | 0.92 |

**Supplementary Table S11. Combined-sample within-person LMM: symptom × group (AFF vs. CTR) interaction.** For each CAP metric (dwell time, DUR; entries, ENT) × symptom (IDS, BAI, FQ) cell, a single LMM was fitted to the combined AFF + CTR sample: rank(CAP) ~ (s_b + s_w) × group + age + sex + site + education level + drug use + (1 | subject), with s_b and s_w z-scored over the combined sample and group CTR-referenced. β s_w CTR and β s_w AFF are the within-person coupling coefficients implied for each group; Δβ (AFF−CTR) is the s_w × group interaction coefficient — the formal test of whether within-person coupling differs between groups — with its SE, t-statistic, Satterthwaite df, raw p-value, and partial eta-squared (η²p = t²/(t² + df)). q is the Benjamini–Hochberg FDR-corrected p-value, applied across the 4 CAPs × 3 symptoms (12 tests) within each metric. Bold indicates q < 0.05. No CAP–symptom pair showed a significant group difference in within-person coupling after correction. N obs / N subj / N AFF / N CTR give the analytic sample for that cell; FQ cells have a smaller N because fear (fearqtsc) was not collected at every session. Diagnostics for this model are reported in Supplementary Tables S10a–S10b.

| **CAP** | **Symptom** | **F** | **df1** | **df2** | ***p*** | ***η²p*** | ***q*** |
| --- | --- | --- | --- | --- | --- | --- | --- |
| **CAP DWELL TIME** | | | | | | | |
| AT-SM | IDS | 1.91 | 2 | 154.00 | 0.15 | 0.02 | 0.90 |
| DM | IDS | 1.96 | 2 | 133.50 | 0.15 | 0.03 | 0.90 |
| FP-DM | IDS | 0.67 | 2 | 162.70 | 0.51 | 0.01 | 0.90 |
| VS-SC | IDS | 0.64 | 2 | 135.70 | 0.53 | 0.01 | 0.90 |
| AT-SM | BAI | 0.29 | 2 | 145.60 | 0.75 | 0.00 | 0.90 |
| DM | BAI | 0.3 | 2 | 130.20 | 0.74 | 0.00 | 0.90 |
| FP-DM | BAI | 0.1 | 2 | 153.30 | 0.90 | 0.00 | 0.91 |
| VS-SC | BAI | 0.09 | 2 | 130.20 | 0.91 | 0.00 | 0.91 |
| AT-SM | FQ | 0.97 | 2 | 135.30 | 0.38 | 0.01 | 0.90 |
| DM | FQ | 0.67 | 2 | 123.10 | 0.51 | 0.01 | 0.90 |
| FP-DM | FQ | 0.77 | 2 | 139.40 | 0.46 | 0.01 | 0.90 |
| VS-SC | FQ | 0.36 | 2 | 123.10 | 0.70 | 0.01 | 0.90 |
| **CAP ENTRIES** | | | | | | | |
| AT-SM | IDS | 0.06 | 2 | 138.50 | 0.95 | 0.00 | 0.95 |
| DM | IDS | 0.68 | 2 | 133.50 | 0.51 | 0.01 | 0.95 |
| FP-DM | IDS | 1.18 | 2 | 135.90 | 0.31 | 0.02 | 0.84 |
| VS-SC | IDS | 1.57 | 2 | 140.70 | 0.21 | 0.02 | 0.84 |
| AT-SM | BAI | 0.24 | 2 | 130.50 | 0.79 | 0.00 | 0.95 |
| DM | BAI | 1.05 | 2 | 130.20 | 0.35 | 0.02 | 0.84 |
| FP-DM | BAI | 0.49 | 2 | 135.90 | 0.61 | 0.01 | 0.95 |
| VS-SC | BAI | 0.24 | 2 | 132.70 | 0.79 | 0.00 | 0.95 |
| AT-SM | FQ | 0.08 | 2 | 127.90 | 0.92 | 0.00 | 0.95 |
| DM | FQ | 0.39 | 2 | 123.10 | 0.68 | 0.01 | 0.95 |
| FP-DM | FQ | 1.7 | 2 | 123.30 | 0.19 | 0.03 | 0.84 |
| VS-SC | FQ | 1.45 | 2 | 123.10 | 0.24 | 0.02 | 0.84 |

**Supplementary Table S12a. Session-moderated within-person LMM, AFF group: omnibus symptom × session (Y0/Y2/Y9) interaction.** For each CAP metric (dwell time, DUR; entries, ENT) × symptom (IDS, BAI, FQ) cell, a within-person LMM was fitted within the AFF group: *rank(CAP) ~ s_b + s_w × session + age + sex + site + education level + drug use + (1 | subject),* with session as a three-level factor (Y0, Y2, Y9) and *s_b, s_w* z-scored within the AFF sample. The reported statistic is the omnibus Kenward-Roger F-test on the full s_w × session interaction, testing whether within-person coupling strengthens or weakens across the nine-year follow-up. q is the Benjamini–Hochberg FDR-corrected p-value, applied across the 4 CAPs × 3 symptoms (12 tests) within the AFF group (a family independent of Table S12b/CTR). Bold indicates q < 0.05. No CAP–symptom pair showed a significant session-moderated interaction after correction (all q ≥ 0.84). Post-hoc pairwise interval contrasts (Y0→Y2, Y0→Y9, Y2→Y9) are reported separately and are not part of this FDR family. Only subjects with ≥ 2 available sessions were retained.

| **CAP** | **Symptom** | **F** | **df1** | **df2** | ***p*** | ***η²p*** | ***q*** |
| --- | --- | --- | --- | --- | --- | --- | --- |
| **CAP DWELL TIME** | | | | | | | |
| AT-SM | IDS | 0.24 | 2 | 144.3 | 0.78 | 0.00 | 0.78 |
| DM | IDS | 0.63 | 2 | 127.2 | 0.53 | 0.01 | 0.65 |
| FP-DM | IDS | 0.42 | 2 | 148.1 | 0.66 | 0.01 | 0.72 |
| VS-SC | IDS | 0.62 | 2 | 121 | 0.54 | 0.01 | 0.65 |
| AT-SM | BAI | 0.69 | 2 | 123.9 | 0.51 | 0.01 | 0.65 |
| DM | BAI | 0.88 | 2 | 113.4 | 0.42 | 0.02 | 0.65 |
| FP-DM | BAI | 0.62 | 2 | 132.4 | 0.54 | 0.01 | 0.65 |
| VS-SC | BAI | 3.8 | 2 | 113.2 | 0.03 | 0.06 | 0.27 |
| AT-SM | FQ | 2.41 | 2 | 150.1 | 0.09 | 0.03 | 0.37 |
| DM | FQ | 1.98 | 2 | 128.5 | 0.14 | 0.03 | 0.43 |
| FP-DM | FQ | 0.68 | 2 | 148.8 | 0.51 | 0.01 | 0.65 |
| VS-SC | FQ | 3.18 | 2 | 128.5 | 0.05 | 0.05 | 0.27 |
| **CAP ENTRIES** | | | | | | | |
| AT-SM | IDS | 1.21 | 2 | 121 | 0.30 | 0.02 | 0.52 |
| DM | IDS | 0.05 | 2 | 131.1 | 0.95 | 0.00 | 1.00 |
| FP-DM | IDS | 1.33 | 2 | 139.5 | 0.27 | 0.02 | 0.52 |
| VS-SC | IDS | 0.12 | 2 | 137.1 | 0.89 | 0.00 | 1.00 |
| AT-SM | BAI | 2.06 | 2 | 115 | 0.13 | 0.03 | 0.40 |
| DM | BAI | 2.21 | 2 | 116.5 | 0.11 | 0.04 | 0.40 |
| FP-DM | BAI | 1.21 | 2 | 121.1 | 0.30 | 0.02 | 0.52 |
| VS-SC | BAI | 2.74 | 2 | 113.2 | 0.07 | 0.05 | 0.40 |
| AT-SM | FQ | 0.22 | 2 | 128.5 | 0.80 | 0.00 | 1.00 |
| DM | FQ | 0.12 | 2 | 138.3 | 0.89 | 0.00 | 1.00 |
| FP-DM | FQ | 3.66 | 2 | 135.3 | 0.03 | 0.05 | 0.34 |
| VS-SC | FQ | 0 | 2 | 143.7 | 1.00 | 0.00 | 1.00 |

**Supplementary Table S12b. Session-moderated within-person LMM, CTR group: omnibus symptom × session (Y0/Y2/Y9) interaction.** Same model and FDR structure as Table S12a, fitted within the CTR group. q is corrected across the 4 CAPs × 3 symptoms (12 tests) within CTR, independently of the AFF family in Table S12a. Bold indicates q < 0.05. No CAP–symptom pair showed a significant session-moderated interaction after correction (all q ≥ 0.27), despite two nominally significant raw p-values that did not survive FDR correction (VS-SC × BAI, DUR: p = 0.03, q = 0.27; FP-DM × FQ, ENT: p = 0.03, q = 0.34). N obs / N subj give the analytic sample for that cell; N Y0/Y2/Y9 give the number of observations contributing to each session. Only subjects with ≥ 2 available sessions were retained.

|  |  |  | **UNADJUSTED** | | | | **ADJUSTED** | | | | |
| --- | --- | --- | --- | --- | --- | --- | --- | --- | --- | --- | --- |
| **Group** | **Interval** | **n** | **rho** | **95% CI** | ***p*** | ***q*** | | **rho** | **95% CI** | ***p*** | ***q*** |
| **ΔFP-DM_CAP_ – ΔDEPRESSION** | | | | | | | | | | | |
| AFF | y0–y2 | 82 | 0.02 | -0.23, 0.20 | 0.89 | 0.96 | | 0.02 | -0.19, 0.24 | 0.83 | 0.98 |
|  | y0–y9 | 49 | 0.07 | -0.34, 0.22 | 0.65 | 0.95 | | 0.05 | -0.33, 0.24 | 0.74 | 0.98 |
|  | y2–y9 | 39 | 0.50 | 0.22, 0.71 | <0.01 | 0.04 | | 0.47 | 0.18, 0.68 | <0.01 | <0.05 |
| CTR | y0–y2 | 34 | 0.09 | -0.42, 0.25 | 0.60 | 0.98 | | 0.10 | -0.43, 0.24 | 0.56 | 0.91 |
|  | y0–y9 | 18 | 0.02 | -0.48, 0.45 | 0.93 | 0.99 | | 0.01 | -0.46, 0.48 | 0.97 | 0.99 |
|  | y2–y9 | 21 | 0.05 | -0.39, 0.47 | 0.82 | 0.99 | | 0.10 | -0.34, 0.51 | 0.65 | 0.91 |
| **ΔVS-SC_CAP_ – ΔFEAR** | | | | | | | | | | | |
| AFF | y0–y2 | 82 | 0.09 | -0.30, 0.13 | 0.42 | 0.95 | | 0.08 | -0.29, 0.14 | 0.49 | 0.98 |
|  | y0–y9 | 49 | 0.36 | 0.09, 0.58 | 0.01 | 0.19 | | 0.44 | 0.18, 0.64 | <0.01 | <0.05 |
|  | y2–y9 | 39 | 0.23 | -0.51, 0.09 | 0.16 | 0.95 | | 0.14 | -0.44, 0.18 | 0.39 | 0.98 |
| CTR | y0–y2 | 34 | 0.09 | -0.41, 0.26 | 0.63 | 0.98 | | 0.21 | -0.51, 0.14 | 0.23 | 0.82 |
|  | y0–y9 | 18 | 0.25 | -0.24, 0.64 | 0.31 | 0.98 | | 0.15 | -0.34, 0.58 | 0.55 | 0.91 |
|  | y2–y9 | 21 | 0.47 | 0.05, 0.75 | 0.03 | 0.98 | | 0.33 | -0.12, 0.67 | 0.14 | 0.82 |
| **ΔAT-SM_CAP_ – ΔFEAR** | | | | | | | | | | | |
| AFF | y0–y2 | 82 | 0.05 | -0.17, 0.26 | 0.67 | 0.96 | | 0.04 | -0.18, 0.25 | 0.74 | 0.97 |
|  | y0–y9 | 49 | 0.31 | -0.54, −0.03 | 0.03 | 0.28 | | 0.37 | -0.59, −0.10 | 0.01 | 0.30 |
|  | y2–y9 | 39 | 0.38 | 0.07, 0.62 | 0.02 | 0.28 | | 0.32 | 0.00, 0.57 | 0.05 | 0.46 |
| CTR | y0–y2 | 34 | 0.13 | -0.22, 0.45 | 0.47 | 0.94 | | 0.22 | -0.13, 0.52 | 0.21 | 0.68 |
|  | y0–y9 | 18 | 0.06 | -0.42, 0.51 | 0.81 | 0.94 | | 0.09 | -0.40, 0.53 | 0.73 | 0.96 |
|  | y2–y9 | 21 | 0.17 | -0.56, 0.28 | 0.45 | 0.94 | | 0.14 | -0.54, 0.31 | 0.55 | 0.96 |

**Supplementary Table S13. Change-score correlations between changes in co-activation pattern metrics and symptom measures.** Spearman correlations (rho) are shown for within-person associations between longitudinal changes in CAP metrics and changes in symptom severity across three assessment intervals (years 0→2, 0→9, and 2→9), separately for cases and controls. Unadjusted and fully adjusted models are presented. The fully adjusted model included age, sex, drug use, scan site, and education level assessed at the start of each interval. *q* values are false discovery rate-adjusted p values computed within each group, interval, and adjustment set, across CAP metrics and symptom measures. n denotes the number of participants with complete data at both time points of the given interval. Abbreviations: CAP, co-activation pattern; CI, 95% confidence interval; FQ, Fear Questionnaire; IDS, Inventory of Depressive Symptomatology; 0, baseline (year 0); 2, 2-year follow-up; 9, 9-year follow-up.
